# The Use of the Integrated Cognitive Assessment (ICA) to Improve the Efficiency of Primary Care Referrals to Memory Services in the Accelerating Dementia Pathway Technologies (ADePT) Study

**DOI:** 10.1101/2023.05.31.23290793

**Authors:** Mohammad Hadi Modarres, Chris Kalafatis, Panos Apostolou, Naji Tabet, Seyed-Mahdi Khaligh-Razavi

## Abstract

**Background:** Current primary care cognitive assessment tools are either crude or time-consuming instruments that can only detect cognitive impairment when it is well established. This leads to unnecessary or late referrals to memory services, by which time the disease may have already progressed into more severe stages.

Due to the COVID-19 pandemic, some memory services have adapted to the new environment by shifting to remote assessments of patients to meet service user demand. However, the use of remote cognitive assessments has been inconsistent, and there has been little evaluation of the outcome of such a change in clinical practice.

Emerging research has highlighted computerised cognitive tests, such as the Integrated Cognitive Assessment (ICA), as the leading candidates for adoption in clinical practice. This is true both during the pandemic and in the post-COVID-19 era as part of healthcare innovation.

**Objectives:** The Accelerating Dementias Pathways Technologies (ADePT) Study was initiated in order to address this challenge and develop a real-world evidence basis to support the adoption of ICA as an inexpensive screening tool for the detection of cognitive impairment and improving the efficiency of the dementia care pathway.

**Methods:** Ninety-nine patients aged 55-90 who have been referred to a memory clinic by a general practitioner (GP) were recruited. Participants completed the ICA either at home or in the clinic along with medical history and usability questionnaires. The GP referral and ICA outcome were compared with the specialist diagnosis obtained at the memory clinic.

Participants were given the option to carry out a retest visit where they were again given the chance to take the ICA test either remotely or face-to-face.

**Results:** The primary outcome of the study compared GP referral with specialist diagnosis of MCI/dementia. Of those the GP referred to memory clinics, 78% were necessary referrals, with ∼22% unnecessary referrals, or patients who should have been referred to other services as they had disorders other than MCI/dementia. In the same population the ICA was able to correctly identify cognitive impairment in ∼90% of patients, with approximately 9% of patients being false negatives. From the subset of unnecessary GP referrals, the ICA classified ∼72% of those as not having cognitive impairment, suggesting that these unnecessary referrals may not have been made if the ICA was in use.

**Conclusions:** The results from this study demonstrate the potential of the ICA as a screening tool, which can be used to support accurate referrals from primary care settings, along with the work conducted in memory clinics and in secondary care.

## Introduction

World-wide, national dementia strategies emphasise the need for improving the diagnostic pathway at the point of primary care towards timely diagnosis. Currently, General Practitioners (GPs) clinical judgement of cognitive impairment is the basis of referral initiation to specialist services. Existing primary care cognitive assessment tools (GPCOG, Mini-Cog, 6CIT etc.), are crude or time-consuming, screening instruments which can only detect cognitive impairment when it is well established. Dementia is difficult to diagnose; in a study concerning false positive diagnoses, 60% of GPs misdiagnosed dementia (Shinagawa, 2016). More detailed tests deployed in secondary-care are expensive and often physically and psychologically intrusive for the patient (e.g. lumbar puncture). As a result many false-positives are identified in referred patients. A key limitation of existing screening tests is the lack of robust evidence to support them; few have been well validated in the populations which they are intended for.

Figure 1 demonstrates the dementia diagnostic pathway for patients. Patients who are referred by GP are triaged. At the memory clinic patients usually undergo two appointments; the first is typically conducted by a nurse or other non-medical professional and involves administration of a cognitive assessment. At the second appointment (i.e. the diagnostic clinic visit), conducted by a dementia medical specialist, the patient receives the outcome of the assessment (see “Outcomes” within Figure 1 for examples of typical outcomes). In light of Covid-19, memory clinics have adapted to the new environment by moving to remote patient assessments in order to continue meeting service user demand while reducing viral transmission. As a result, the majority of appointments were conducted remotely with the use of pen-and-paper tests that are acceptable for remote use.

**Figure 1.**
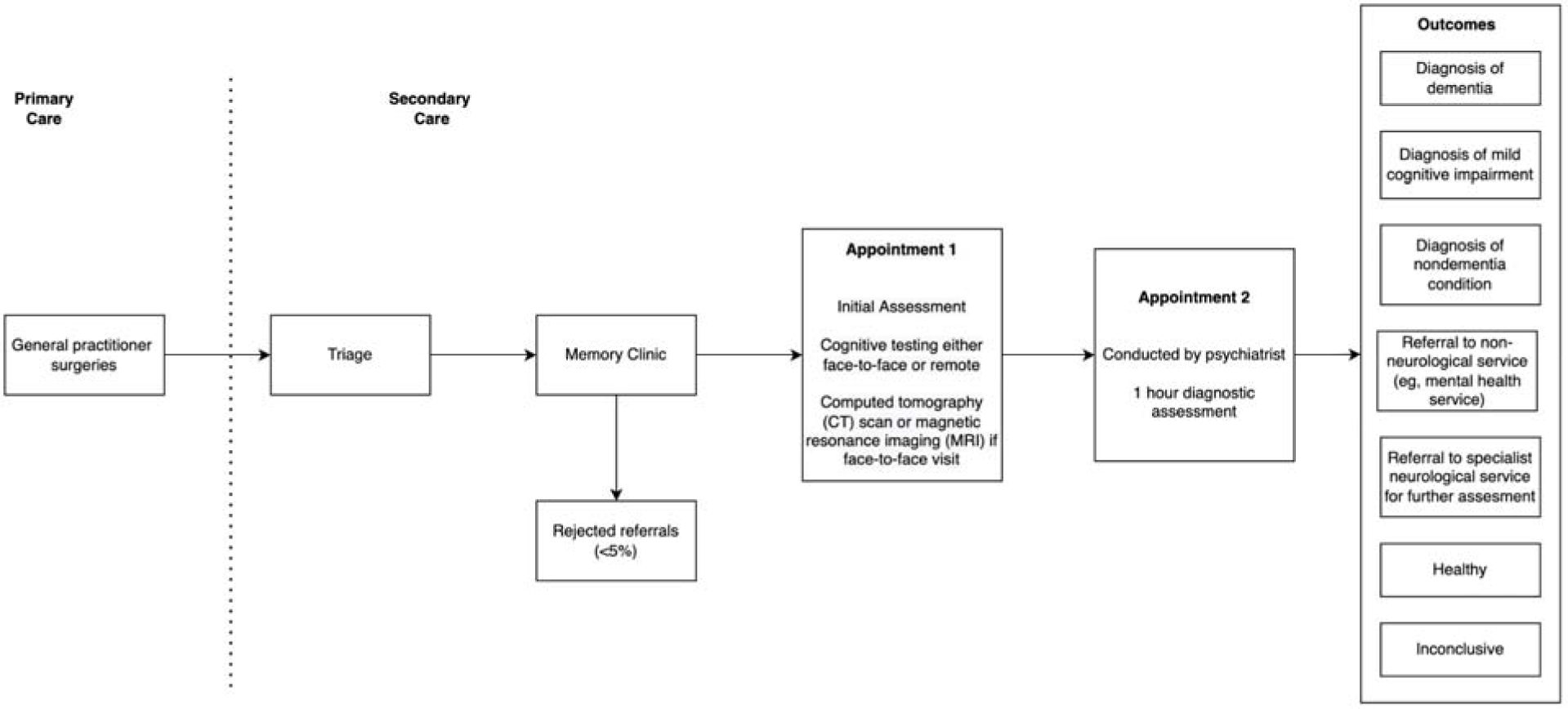
Memory clinic pathway.

The COVID-19 pandemic effectively brought clinical practice in the memory services to a standstill. Nationally, memory services adapted to the new environment by moving to remote patient assessments in order to continue meeting service user demand while reducing viral transmission (Owens at al. 2020). However, the remote use of cognitive assessments has been variable, non-standardised while there has been scant evaluation of the outcome of such a change in clinical practice (Binng et al. 2020). Emerging research in remote memory clinics has highlighted computerised cognitive tests such as the ICA as a prominent candidate for adoption in clinical practice both during the pandemic and for post-COVID implementation as part of healthcare innovation (Dunne at al. 2020).

The ICA is a 5-minute, self-administered computerised cognitive test based on a rapid categorisation task that employs an Artificial Intelligence model to improve its accuracy in detecting cognitive impairment (Kalafatis et al., 2021). The ICA is self-administered and independent of language (Khaligh-Razavi et al., 2019, Khaligh-Razavi et al., 2020). The value proposition of the ICA is that a more accurate and sensitive tool for diagnosis will streamline the diagnosis of dementia by reducing false positive results from GP referrals and therefore minimising the need for further, expensive and time-consuming assessments.

The Accelerating Dementias Pathways Technologies (ADePT) Study was initiated in order to address this challenge and develop a real-world evidence basis to support the adoption of ICA as an inexpensive screening tool for the detection of cognitive impairment and improving the efficiency of the dementia care pathway.

The primary objective for the ADePT study was to deliver real-world evidence on practices and the economic case for ICA adoption in memory clinics for the assessment of cognitive impairment associated with dementia, Alzheimer’s Disease (AD), Mild Cognitive Impairment (MCI) and similar diseases, including assessment of preferred business models by comparing the precision of GP referrals against the ICA.

This report focuses on the results from the clinical study, which is one of the work packages of the ADePT study. The results from this study inform the health economics model (e.g. Shore et al, 2022), which has been delivered separately by York Health Economics Consortium.

## Methods

### The ICA Test Description

The ICA test has been described in detail in previous publications (Khaligh-Razavi et al. 2019, Kalafatis et al. 2021). The ICA test is a rapid visual categorisation task. The test takes advantage of the human brain’s strong reaction to animal stimuli. One hundred natural images (50 of animals and 50 of not containing an animal) of various levels of difficulty are selected and are presented to the participant in rapid succession.

Each image is presented for 100 ms followed by a 20 ms inter-stimulus interval (ISI), followed by a dynamic noise mask (for 250 ms), followed by the subject’s categorisation into animal vs. non-animal.

### Accuracy, Speed and Summary ICA Index Calculation

The raw data from the ICA is composed of reaction time and categorisation accuracy on the images. This data was used to calculate summary features such as overall accuracy, and speed using the same methodology as described previously (Khaligh-Razavi et al. 2019, Kalafatis et al. 2021).

Accuracy is defined as follows:

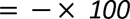

Speed is defined based on participant’s response reaction times in trials they responded correctly:

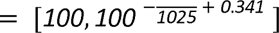

A summary ICA Index, is calculated as follows:

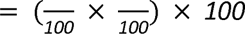

The ICA Index describes the raw test result, incorporating speed and accuracy, the two main elements of the ICA test.

### ICA AI Model

The AI model utilises inputs from accuracy and speed of responses to the ICA rapid categorisation task (with the ICA Index as an input feature), as well as age, and outputs an indication of likelihood of impairment (AI probability) by comparing the test performance and age of a patient to those previously taken by healthy and cognitively impaired individuals. The AI model is able to achieve an improved classification accuracy relative to using any single feature from the ICA test.

A probability threshold value of 0.5 was used to convert the AI probability to the AI prediction of healthy or cognitively impaired (MCI/mild AD). The AI probability was also converted to a score between 0 and 100 using the following equation:

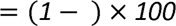

The ICA AI model used in this study was a binary logistic regression machine learning model which is a supervised linear classifier. The algorithm’s task is to learn a set of weights from a regression model that maps the participant’s ICA test results and demographics to the classification label of healthy or cognitively impaired.

### Ethics Approval

Health Research Authority and Health and Care Research Wales approval for this study was obtained in February 2020. The study is registered in the ISRCTN Registry (ISRCTN16596456).

### Study Design

All participants were recruited among attendees at the National Health Service (NHS) memory assessment services at the point of referral by their GP. Participants were recruited from Devon Partnership NHS Trust, North Bristol NHS Foundation Trust, Oxford Health NHS Foundation Trust, and Sussex Partnership NHS Foundation Trust.

The participants who did not have a formal diagnosis of a neurodegenerative disease were triaged as per usual clinical practice and were asked to complete the ICA in parallel with the diagnostic assessment.

The main study inclusion criterion was referral to the memory clinic by a GP. Patients recruited were 55 to 90 years old. Potential participants had to be fully informed of and understand the objectives, procedures, and possible benefits and risks of the study and have the capacity to provide written consent.

Subjects that met the following criteria were excluded from the study cohort:

- Lack of capacity to consent to participation in this study
- Upper limb arthropathy or motor dysfunction that limits the use of a tablet computer
- Visual impairment severe enough to limit the use of a tablet computer
- Known diagnosis of dementia
- Already receiving cholinesterase inhibitors and/or Memantine

### Study Procedures

Participants enrolled in the study were required to attend one visit at a designated memory clinic or remotely at their home (Assessment Visit 1 [AV1]). Participants were asked to complete the ICA. Prior to taking the ICA, participants were requested to view a short training video to assist them in completing the task successfully. After taking the ICA, patients completed the following short questionnaires:

- Inquiry on stimulants, fatigue, and sleep: A questionnaire that assesses the participant’s overall state. Questions revolve around recent intake of stimulants (eg, coffee or alcohol), sleep quality, energy levels, and mood. The questionnaire was used in conjunction with the ICA to determine whether any of these factors have had an impact on ICA performance.
- ICA Usability Questionnaire: A questionnaire that assesses the participant’s views on their experience with the test to receive acceptability and usability feedback for the ICA.
- Cognitive Health Questionnaire: A questionnaire that assesses the participant’s history of activities of daily living and physical and mental health comorbidities. The questions should ideally be answered by the informant (study partner) if available or by the participant if an informant is not present. The questionnaire was used in conjunction with the ICA to determine whether cognitive impairment detected by the ICA was due to MCI/dementia or other organic and/or treatable conditions. The results from the cognitive health questionnaire are shown in Appendix A3.

Lastly, a brief medical history of the participants via electronic health care records was obtained, mainly focusing on any cognitive tests that have been taken by the participants.

Participants were given the option to carry out a retest visit (Assessment Visit 2 [AV2]) whereby they were again given the chance to take the ICA test either remotely or face-to-face, complete a usability questionnaire, and respond to inquiries on stimulants, fatigue, and sleep. The overall study pathway for participants is detailed at a high level within Figure 2.

**Figure 2.**
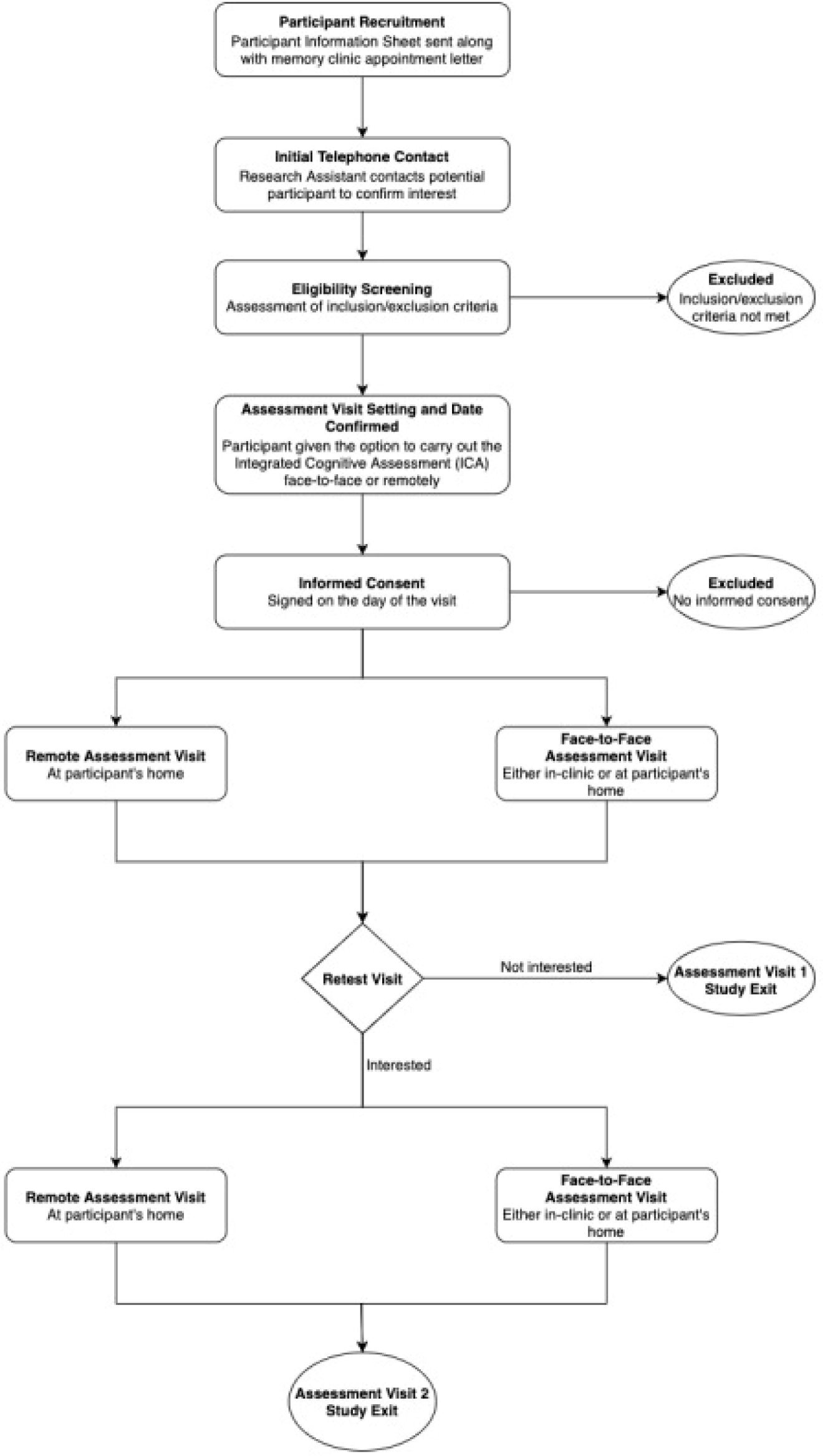
Accelerating Dementia Pathway Technologies (ADePT) study participant pathway.

### Statistical Analysis

For the purposes of these analyses, patients referred to the memory clinic were divided into the following 3 groups, based on their memory clinic outcome: (A) those who receive a diagnosis of MCI or dementia, (B) those who are identified as healthy or receive a diagnosis of a brain or mental disorder other than MCI or dementia, and (C) those who receive an inconclusive diagnosis.

Participants with an inconclusive outcome after the memory clinic assessment were excluded from further analysis.

Participants in group A were counted as correct GP referrals. Participants in group B were counted as unnecessary or incorrect referrals.

Comparison with Specialist Diagnosis of MCI/Dementia The metrics for GP referrals that were calculated are the following:

- Total number of patients referred by GPs=A+B+C
- Proportion of necessary GP referrals (excluding inconclusive)=A/(A+B)
- Proportion of unnecessary GP referrals (excluding inconclusive)=B/(A+B)

Likewise, the following complementary metrics for the ICA were calculated:

- Total number of patients the ICA would have referred l1 Proportion of patients correctly referred by the ICA
- Proportion of patients incorrectly referred by the ICA
- Proportion of patients correctly not referred by the ICA
- Proportion of patients incorrectly not referred by the ICA

In a secondary outcome analysis, we compared with specialist diagnosis of all types of cognitive impairment (those due to MCI, dementia, or other neurological or mental disorders).

### Comparison with other cognitive tests

Cut-offs were applied to cognitive tests taken in primary care and secondary care, to investigate the predictive accuracy of these tests compared to the clinical diagnosis determined at the memory clinic. Cognitive tests taken in the 6 months prior to the memory clinic assessment were used for the analysis. The results from the cognitive tests are compared to ICA results on the same patients. The cognitive tests used for this analysis were ACE-III, MoCA, MoCA Blind, GPCOG and SCIT. A sufficient number of participants had completed these assessments within 6 months of the memory clinic visit to make the analysis viable.

We calculated the correlation between ICA Index with cognitive tests taken in memory clinics and GP settings where there was at most 12 months difference in date between the tests. Results were included where there were at least 10 data points for the correlation calculation (see Appendix A1 for results).

### Test-Retest Analysis

The test-retest reliability of the ICA was analysed by the following:

- Calculation of intraclass correlation coefficient to assess test-retest reliability across all participants
- Scatterplot construction and calculation of correlation coefficient between the initial and final assessment for all participants
- Construction of Bland-Altman plots for the initial and final assessment to assess agreement

### Qualitative Data from Usability Questionnaire

Multiple choice responses from participants were analysed by calculating the proportion of participants who selected each option. Questions relating to frequency of tablet or mobile phone use were used to assess familiarity with technology, in particular touch screen devices. The ease of understanding the ICA instructions and level of difficulty of the categorization task was analysed by calculating the proportion of participants who reported finding each of these steps very easy, easy, moderately difficult, difficult, or very difficult.

### Procedures to Account for Missing and Spurious Data

Patients with inconclusive outcomes were excluded from our analysis. Other than that we did not have any other missing data regarding the calculations needed for primary and secondary outcome measures.

## Results

### Participants

Of a total of 99 participants, 86 participants completed all assessments and questionnaires successfully. Some participants did not complete the ICA (13.13%), the majority of these were patients diagnosed with dementia (69.2%), followed by 7.7% of patients with MCI, 7.7% inconclusive, 7.7% non-dementia conditioned and 7.7% healthy. Of those who did not complete the trial, 11 did not complete the ICA, either due to inability to complete the test, or due to the Researcher’s decision to cancel the test while 1 participant decided to not proceed with the study procedures. Finally there was also 1 participant that was discontinued due to passing away prior to receiving their diagnosis. Table 1 illustrates the recruited participants breakdown.

**Table 1.** Recruited participants by diagnosis following reclassification. Recruitment is broken down by face to face and remote visits, and whether or not the participant completed all assessments including the ICA.

| Diagnosis | Completed ICA |  |  | Did not complete |  |  | Overall |  |
| --- | --- | --- | --- | --- | --- | --- | --- | --- |
|  | Face to Face | Remote | Total | Face to Face | Remote | Total | Overall total | Percent completed |
| Dementia | 35 | 10 | 45 | 8 | 1 | 9 | 54 | 83.3 |
| MCI | 13 | 5 | 18 | 1 | 0 | 1 | 19 | 94.7 |
| Healthy | 10 | 5 | 15 | 1 | 0 | 1 | 16 | 93.3 |
| Inconclusive | 4 | 1 | 5 | 1 | 0 | 1 | 6 | 85.7 |
| Non-dementia condition | 3 | 0 | 3 | 1 | 0 | 1 | 4 | 75.0 |
| Total | 65 | 21 | 86 | 12 | 1 | 13 | 99 | 86.9 |

The distribution of age and education years for all participants is shown in Table 2, and broken down by diagnosis in Table 3.

**Table 2.**
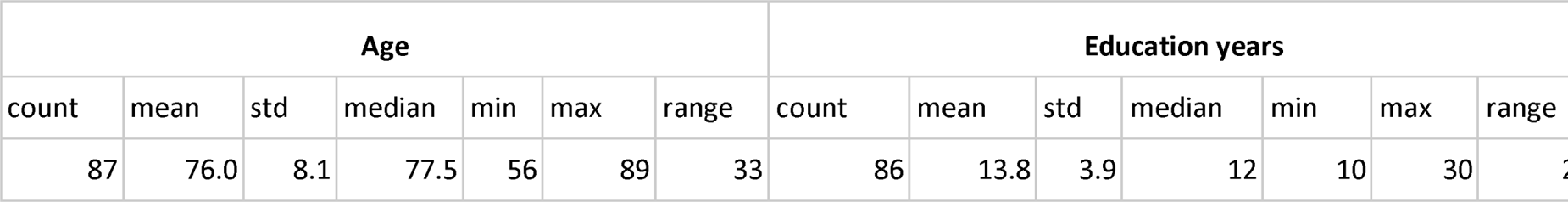
Age and education years of all participants

**Table 3.** Age and education years of participants per diagnosis

| Diagnosis | Age |  |  |  |  |  |  | Education years |  |  |  |  |  |  |
| --- | --- | --- | --- | --- | --- | --- | --- | --- | --- | --- | --- | --- | --- | --- |
|  | count | mean | std | median | min | max | range | count | mean | std | median | min | max | range |
| Dementia | 45 | 79.1 | 6.9 | 79 | 63 | 89 | 26 | 45 | 13.2 | 3.5 | 12 | 10 | 24 | 14 |
| MCI | 18 | 74.7 | 5.5 | 74.5 | 64 | 82 | 18 | 18 | 13.8 | 3.3 | 13 | 10 | 20 | 10 |
| Healthy | 14 | 69.2 | 8.9 | 68.5 | 57 | 85 | 28 | 14 | 15.7 | 5.6 | 15 | 11 | 30 | 19 |
| Inconclusive | 6 | 70.7 | 11.3 | 72.5 | 56 | 85 | 29 | 6 | 14.8 | 4.4 | 13.5 | 11 | 23 | 12 |
| Non-dementia condition | 3 | 78.3 | 5.5 | 81 | 72 | 82 | 10 | 3 | 12.3 | 2.5 | 12 | 10 | 15 | 5 |

### Comparison of GP/ICA referrals with specialist diagnosis

The following analysis is based on participants who completed the required assessments and questionnaires.

### Comparison of GP/ICA referral with specialist diagnosis of MCI/dementia

From the 86 participants who also completed the ICA, 5 also had inconclusive diagnosis, hence the number of participants under investigation for the primary outcome analysis is 81.

**Table 4.** Comparison of precision of GP referrals with specialist diagnosis of MCI/dementia

|  |  |
| --- | --- |
| total referred by GP | 86 |
| total referred by GP excluding inconclusive | 81 |
| those with diagnosis of MCI or dementia | 63 |
| those who are healthy or with diagnosis of disorders other than MCI/dementia | 18 |
| inconclusive diagnosis | 5 |
| proportion of necessary referrals by GP | 77.8 |
| proportion of unnecessary referrals by GP | 22.2 |

**Table 5.** GP Classification matrix

|  |  | GP referral |  |
| --- | --- | --- | --- |
|  |  | Not referred | Referred |
| Specialist diagnosis | Healthy/Other | NA | 18 |
|  | MCI/Dementia | NA | 63 |

**Table 6.** AI V1 Classification matrix

|  |  | ICA AI V1 |  |
| --- | --- | --- | --- |
|  |  | Healthy | Impaired |
| Specialist diagnosis | Healthy/Other | 13 | 5 |
|  | MCI/Dementia | 6 | 57 |

**Table 7.** Comparison of ICA referral with specialist diagnosis of MCI/dementia

|  | Primary outcome Analysis |
| --- | --- |
|  | AI V1 |
| total ICA would have referred | 62 |
| proportion correctly referred by ICA | 90.5 |
| proportion incorrectly not referred by ICA | 9.5 |
| proportion correctly not referred by ICA | 72.2 |
| proportion incorrectly referred by ICA | 27.8 |

### Comparison of GP/ICA precision in referral with specialist diagnosis of all types of impairment

In this analysis the precision of GP referral is compared to specialist diagnosis of MCI, dementia and all other types of non-dementia neurological/mental disorders.

**Table 8.** Comparison of GP referral precision with specialist diagnosis of all types of impairment

|  |  |
| --- | --- |
| total referred by GP | 86 |
| total referred by GP excluding inconclusive | 81 |
| those with diagnosis of MCI or dementia or other neurological/mental disorders | 66 |
| those who are healthy | 15 |
| inconclusive diagnosis | 5 |
| proportion of necessary referrals by GP | 81.5 |
| proportion of unnecessary referrals by GP | 18.5 |

**Table 9.** GP Classification matrix

|  |  | GP referral |  |
| --- | --- | --- | --- |
|  |  | Not referred | Referred |
| Specialist diagnosis | Healthy | NA | 15 |
|  | MCI/Dementia/Other | NA | 66 |

**Table 10.**
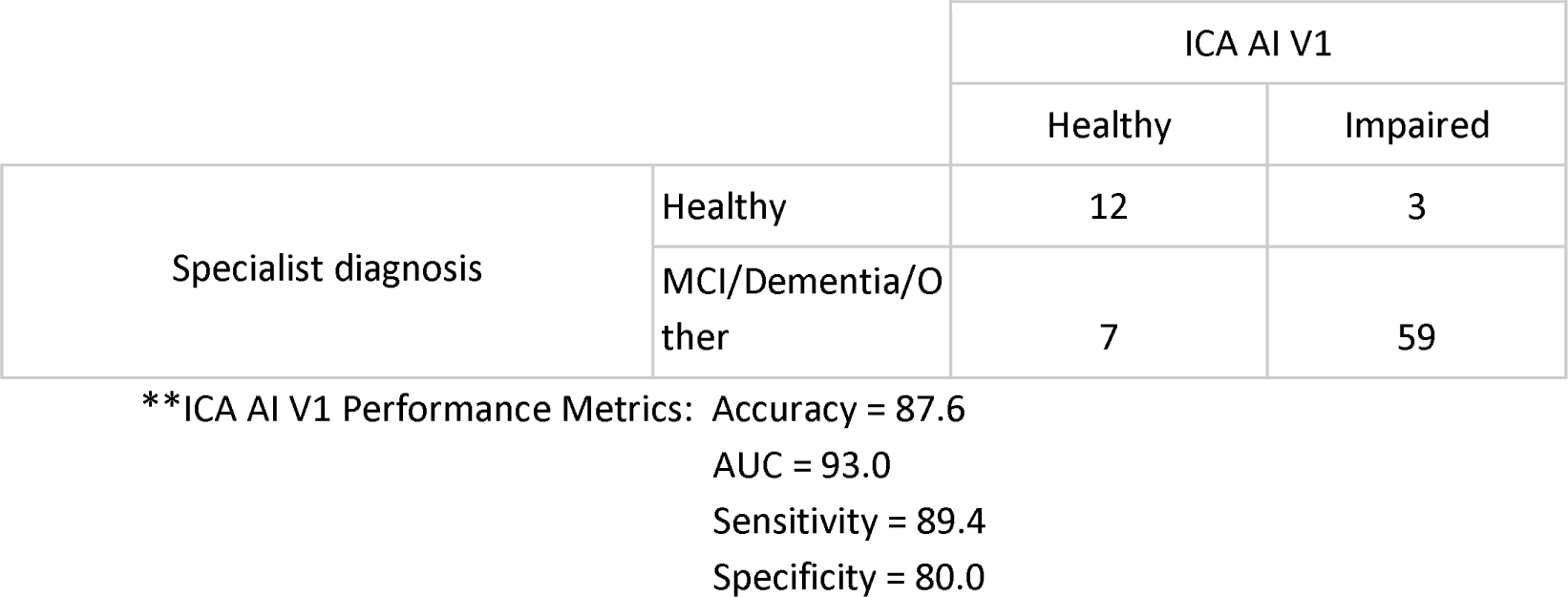
AI V1 Classification matrix and performance metrics (healthy vs impaired).

**Table 11.** Comparison of ICA referral with with specialist diagnosis of all types of impairment

|  | Secondary outcome Analysis |
| --- | --- |
|  | AI V1 |
| total ICA would have referred | 62 |
| proportion correctly referred by ICA | 89.4 |
| proportion incorrectly not referred by ICA | 10.6 |
| proportion correctly not referred by ICA | 80.0 |
| proportion incorrectly referred by ICA | 20.0 |

### Test-retest analysis

There were 24 participants who completed the initial visit and the retest. The days between the test and retest were 9.7 on average (SD of 3 days, minimum of 5 days, maximum of 14 days). The correlation between the ICA test retest is shown in Figure 3. The Spearman Rank coefficient for the test-retest ICA Index was calculated to be 0.79 (p<0.000). While the average ICA Index in the retest group was 3.8 points higher than the initial test (see Bland Altman plot in Figure 10), paired t-test p-value was >0.1, suggesting this was not a statistically significant increase in score on the retest (Table 12). The Spearman Rank coefficient for the test-retest ICA Score (V1) was found to be 0.81 (p<0.000) (Table 12).

**Figure 3.**
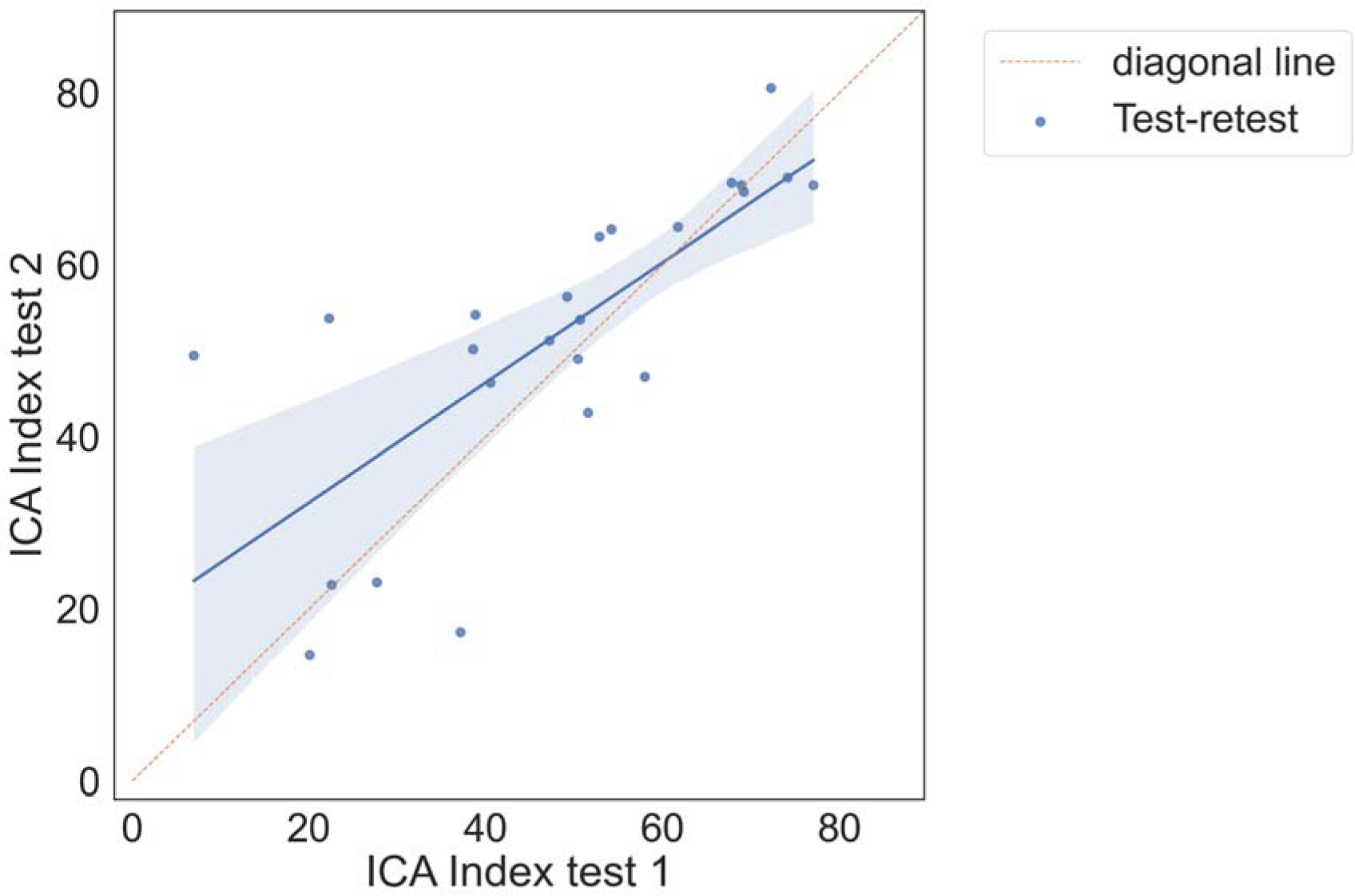
Correlation plot between test and retest, showing a strong positive correlation between the two tests.

**Figure 4.**
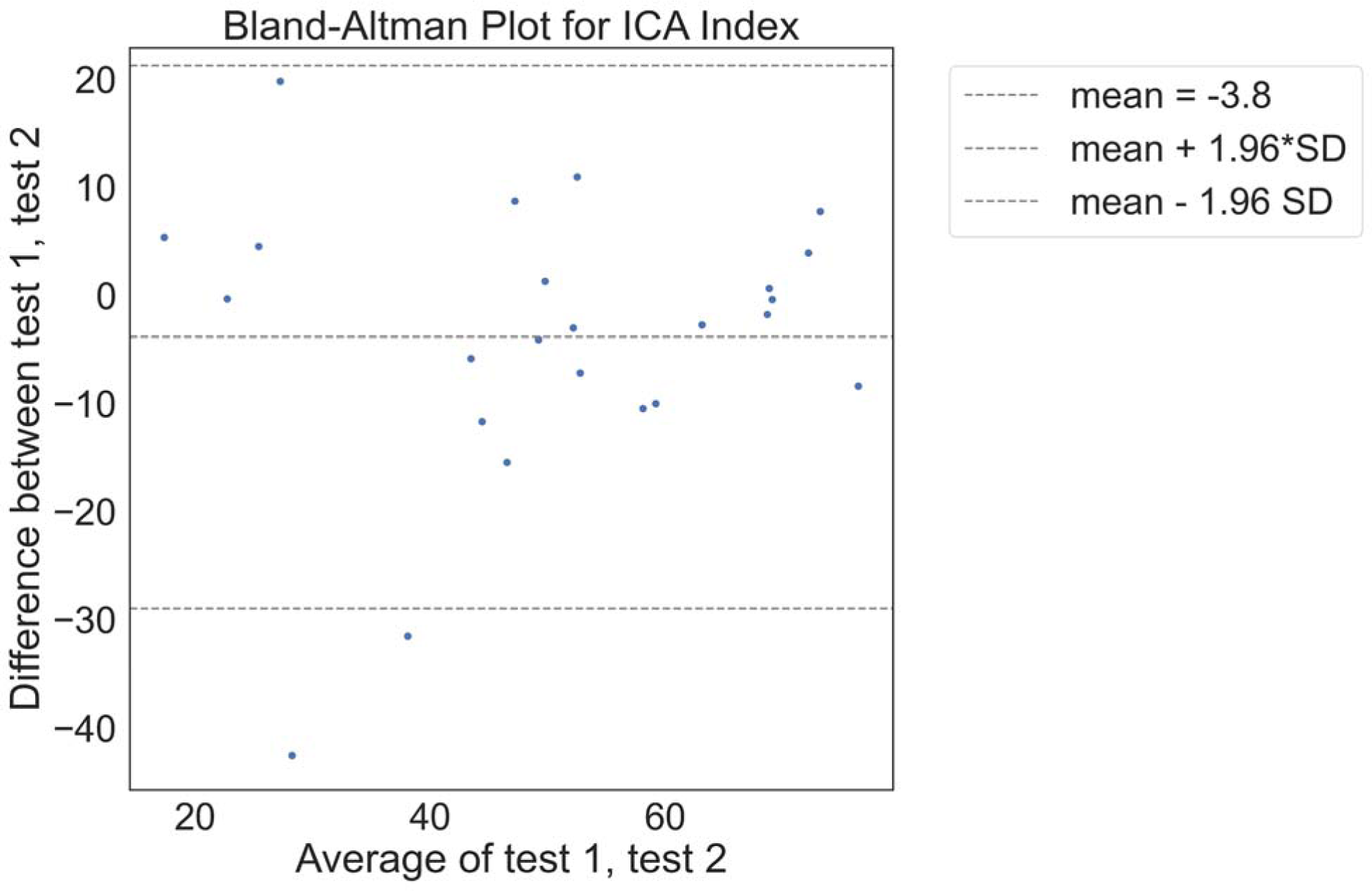
Bland Altman plot of differences between test and retest

**Table 12.** Correlation coefficients and other metrics associated with test-retest

| Metric | ICA Index |
| --- | --- |
| pearson r | 0.75 |
| pearson p-value | 0.00 |
| spearman r | 0.79 |
| spearman p-value | 0.00 |
| ICC | 0.738[0.49 0.88] |
| t-test statistic | -1.43 |
| t-test p-value | 0.17 |
| Cohen's D | -0.21 |
| mean test 1 | 48.32 |
| mean test 2 | 52.14 |
| difference test1-test2 | -3.81 |
| sd test 1 | 18.59 |
| sd test 2 | 17.35 |

The 2×2 matrix of ICA prediction on the first and second test is shown in Table 13.

**Table 13.** ICA prediction on first and second visit.

|  |  | ICA Prediction visit 2 |  |
| --- | --- | --- | --- |
|  |  | Healthy | Impaired |
| ICA prediction visit 1 | Healthy | 7.00 | 0.00 |
|  | Impaired | 3.00 | 14.00 |

The test retest prediction agreement for the ICA is 87.5%.

### Comparison of cognitive test cut-offs with ICA

In this section cut-offs are applied to cognitive tests taken in primary care and secondary care, to investigate the predictive accuracy of these tests compared to the clinical diagnosis determined at the memory clinic. Cognitive tests taken in the 6 months prior to the memory clinic assessment were used for the analysis. The results from the cognitive tests are compared to ICA results on the same patients. The cognitive tests used for this analysis are ACE-III, MoCA, MoCA Blind, GPCOG and SCIT. A sufficient number of participants had completed these assessments within 6 months of the memory clinic visit to make the analysis viable.

### ACE-III memory clinic assessment

The most common memory clinic assessment was ACE-III. This was completed by 68 participants. A cut off score of ≥88 was used for ACE-III scores for binary prediction of healthy and impaired. On average the ACE and ICA assessments were conducted 38 days and 24 days from the diagnosis date, respectively.

ACE outcomes for participants, broken down by diagnosis, are shown in Table 14, and compared with ICA outcomes for the same patients. A comparison of these cognitive tests should also consider that the average ICA completion time is ∼9 mins, compared to approximately 30 mins for ACE-III.

**Table 14.** ACE and ICA labels compared to the clinical diagnosis

|  |  | ACE |  | ICA |  |  |
| --- | --- | --- | --- | --- | --- | --- |
| Clinical diagnosis | Total | Healthy | Impaired | Healthy | Impaired | Did not complete |
| Dementia | 37 | 1 | 36 | 3 | 30 | 4 |
| MCI | 14 | 5 | 9 | 2 | 12 | 0 |
| Inconclusive | 2 | 0 | 2 | 0 | 1 | 0 |
| Healthy | 12 | 12 | 0 | 9 | 2 | 1 |
| Non-dementia condition | 3 | 2 | 1 | 0 | 2 | 1 |
| Total | 68 | 20 | 48 | 14 | 47 | 6 |

Excluding inconclusive cases, the overall accuracy of ACE-III on these participants (n=66) is 88%. Here we assumed that detecting cognitive impairment in non-dementia cases is a correct classification.

Comparing ICA and ACE predictions, where both the ICA and ACE was completed, results in the matrix shown in Table 15.

**Table 15.**
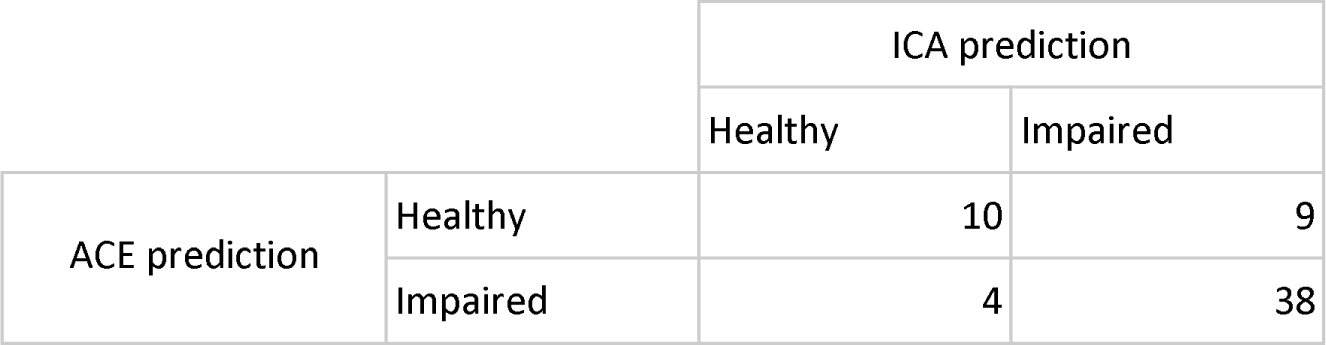
ICA and ACE prediction comparison in 2×2 matrix

|  |  | ICA prediction |  |
| --- | --- | --- | --- |
|  |  | Healthy | Impaired |
| ACE prediction | Healthy | 10 | 9 |
|  | Impaired | 4 | 38 |

The overall percent agreement between the ICA and ACE is 79%. Supplementary Table S3 shows the diagnosis, ICA and ACE scores for the participants where the ICA and ACE had differing predictions.

### MoCA memory clinic assessment

For MoCA assessments, which were conducted in memory clinics, a cut-off of 26 or greater was used to predict healthy patients. On average the ICA test was taken 17 days apart from the diagnosis date, while the MoCA assessment was taken 58 days from diagnosis.

**Table 16.** Comparison of predictions from MoCA and ICA

| Diagnosis | Total | MoCA_Healthy | MoCA_Impaired | ICA_did_not_complete | ICA_Healthy | ICA_Impaired |
| --- | --- | --- | --- | --- | --- | --- |
| Dementia | 3 | 0 | 3 | 2 | 0 | 1 |
| Healthy | 3 | 3 | 0 | 0 | 2 | 1 |
| MCI | 3 | 2 | 1 | 0 | 1 | 2 |
| Inconclusive | 2 | 1 | 1 | 0 | 0 | 2 |
| Non-dementia condition | 1 | 1 | 0 | 0 | 1 | 0 |
| Totals | 12 | 7 | 5 | 2 | 4 | 6 |

Excluding inconclusive cases and non-dementia conditions, the overall accuracy of MoCA on these participants (n=10) is 70%. Here we assumed that detecting cognitive impairment in non-dementia cases is a correct classification.

**Table 17.** 2×2 comparison of MoCA and ICA predictions

|  |  | ICA prediction |  |
| --- | --- | --- | --- |
|  |  | Healthy | Impaired |
| MoCA prediction | Healthy | 4 | 3 |
|  | Impaired | 0 | 3 |

Overall the ICA and MoCA had a percentage agreement of 70%. The cases where the ICA and MoCA prediction were different are shown in supplementary Table S4.

### MoCA blind memory clinic assessment

This analysis was based on MoCA Blind assessments carried out by the memory clinic. A score ≥18 for healthy was used as the cut-off for healthy participants (Mahendran, et al 2015). The ICA assessment was 15 days from diagnosis date, while the MoCA Blind assessment was 16 days from diagnosis date.

**Table 18.** Comparison of predictions from MoCA Blind and ICA

| Diagnosis | Total | MoCA_blind_Healthy | MoCA_blind_Impaired | ICA_did_not_complete | ICA_Healthy | ICA_Impaired |
| --- | --- | --- | --- | --- | --- | --- |
| Dementia | 13 | 0 | 13 | 2 | 0 | 11 |
| MCI | 1 | 1 | 0 | 0 | 0 | 1 |
| Totals | 14 | 1 | 13 | 2 | 0 | 12 |

The overall accuracy of MoCA Blind for these participants (n=14) is 93%. Here we assumed that detecting cognitive impairment in non-dementia cases is a correct classification.

**Table 19.** 2×2 comparison of MoCA Blind and ICA predictions

|  |  | ICA prediction |  |
| --- | --- | --- | --- |
|  |  | Healthy | Impaired |
| MoCA Blind prediction | Healthy | 0 | 1 |
|  | Impaired | 0 | 11 |

The ICA and MoCA Blind had a 93% agreement rate. There was only one instance (from 12) in which the assessments differed (Table S5). In this case the ICA predicted impairment, in agreement with the clinical diagnosis of MCI, while the MoCA Blind score of 18 was suggestive of a healthy patient.

### GPCOG in primary care assessment

The following cut offs were used for the GPCOG assessment, based on (Brodaty et al, 2004):

- Score of 0-4: cognitive impairment indicated
- Score of 5-8: more information required
- Score of 9: no significant impairment (for this case we have indicated Healthy for direct comparison with the ICA)

The ICA test was completed on average 15 days before diagnosis, while the GPCOG test was completed on average 67 days before diagnosis.

**Table 20.** Comparison of predictions from GPCOG and ICA

| clinical_diagnosis_short | Total | GPCOG_Further Investigate | GPCOG_Healthy | GPCOG_Impaired | ICA_did_not_complete | ICA_Healthy | ICA_Impaired |
| --- | --- | --- | --- | --- | --- | --- | --- |
| **Dementia** | 12 | 4 | 3 | 5 | 3 | 2 | 7 |
| **Healthy** | 3 | 0 | 3 | 0 | 0 | 2 | 1 |
| **MCI** | 1 | 0 | 1 | 0 | 0 | 0 | 1 |
| Totals | 16 | 4 | 7 | 5 | 3 | 4 | 9 |

The overall accuracy of GPCOG for these participants (n=16) is 75%. Here we assumed that detecting cognitive impairment in non-dementia cases is a correct classification. We also combined ‘Further Investigate’ with ‘Impaired’ to indicate that GPCOG had identified cognitive impairment.

**Table 21.** Comparison matrix of GPCOG and ICA predictions

|  |  | ICA prediction |  |
| --- | --- | --- | --- |
|  |  | Healthy | Impaired |
| GPCOG prediction | Further Investigate | 2 | 1 |
|  | Healthy | 2 | 5 |
|  | Impaired | 0 | 3 |

### 6CIT in primary care assessment

For the 6CIT assessment, carried out at GPs, a cut off score of 8 or greater was used to indicate cognitive impairment, based on (Brooke and Bullock, 1999). For the participants who completed the 6CIT and ICA, the ICA was completed on average 31 days before diagnosis, while the 6CIT test was completed on average 103 days before diagnosis.

**Table 22.** Comparison of predictions from 6CIT and ICA

| Diagnosis | Total | SCIT_Healthy | SCIT_Impaired | ICA_Healthy | ICA_Impaired |
| --- | --- | --- | --- | --- | --- |
| **Dementia** | 11 | 3 | 8 | 1 | 9 |
| **Healthy** | 2 | 1 | 1 | 2 | 0 |
| **Inconclusive** | 1 | 1 | 0 | 0 | 1 |
| Totals | 14 | 5 | 9 | 3 | 10 |

Excluding inconclusive cases, the overall accuracy of 6CIT for these participants (n=13) is 69%.

**Table 23.** 2×2 comparison of 6CIT and ICA predictions

|  |  | ICA prediction |  |
| --- | --- | --- | --- |
|  |  | Healthy | Impaired |
| SCIT prediction | Healthy | 1 | 4 |
|  | Impaired | 2 | 6 |

Finally, the performance between ICA and other cognitive tests is summarised in the table below:

**Table 24.** Performances by ICA and standard care tests (meta-data) at glance

|  | MCI (AUC=87%) |  | Dementia (AUC=96%) |  |  |
| --- | --- | --- | --- | --- | --- |
|  | Sensitivity | Specificity | Sensitivity | Specificity |  |
| ICA tool | 83% | 80% | 93% | 80% |  |
|  | MCI |  | Dementia |  |  |
| Standard care test* | Sensitivity | Specificity | Sensitivity | Specificity | Proportion of patients receiving |
| Mini-Mental State Examination (MMSE) | 51% | 75% | 59% | 85% | 26% |
| General Practitioner Assessment of Cognition (GPCOG) | 52% | 82% | 60% | 93% | 21% |
| 6-Item Cognitive Impairment Test (6CIT) | 66% | 70% | 88% | 78% | 29% |
| Test Your Memory (TYM) |  |  | 93% | 86% | 0% |
| Memory Impairments Screen (MIS) |  |  | 80% | 91% | 0% |
| Free Cog |  |  |  |  | 0% |
| Telephone interview for Cognitive Status (TICS) |  |  |  |  | 0% |
| Abbreviated mental test score (AMTS) | 66% | 70% | 81% | 84% | 7% |
| Montreal Cognitive Assessment (MoCA) | 80% | 81% | 91% | 81% | 6% |
| Other | 63% | 76% | 79% | 85% | 11% |
| **Weighted** | **60%** | **75%** | **73%** | **84%** |  |

\* See Shore, et al. (2022) for a summary of relevant literature reporting on standard care tests.

## ICA Usability

### ICA completion rate

An update to the CognICA application implemented a new test flow, which automated the pre-test demo/trial image process. More specifically, the pre-test image process was automated while a training video was introduced in order to facilitate remote assessments. These changes did not affect the core of the test.

This led to an increase in the completion rate of the ICA amongst the impaired diagnosis groups (Table 25). For example for dementia patients the completion rate increased from 78% to 89%.

**Table 25.** Completion rate of the ICA, across diagnoses, based on the cognICA version

|  | CognICA version 1.2.1 |  |  |  | CognICA version 1.3.2 |  |  |  | Overall total |
| --- | --- | --- | --- | --- | --- | --- | --- | --- | --- |
| Diagnosis | Completed | Did not complete | Completion (%) | Total | Completed | Did not complete | Completion (%) | Total |  |
| **Dementia** | 21 | 6 | 77.8 | 27 | 24 | 3 | 88.9 | 27 | 54 |
| **MCI** | 6 | 1 | 85.7 | 7 | 12 | 0 | 100.0 | 12 | 19 |
| **Healthy** | 5 | 0 | 100.0 | 5 | 10 | 1 | 90.9 | 11 | 16 |
| **Inconclusive** | 2 | 0 | 100.0 | 2 | 4 | 0 | 100.0 | 4 | 6 |

A further benefit of the updated test flow was a reduction in the number of trial images completed by patients. The completion rate of the ICA could also have been influenced by the new training video. This was introduced partway through the study, and is discussed in more detail further on. However, amongst dementia patients, there was an increased rate of completion (from 80% to 85%) when the video was watched compared to when it was not watched.

### ICA completion time

On average, from device hand over to completion of the test the ICA took ∼9 minutes to complete in this setting. This is based on the researcher’s approximation of the time taken, and is not a precise measurement. The mean time taken for ICA completion was reduced from visit 1 to visit 2, from 9 min to 7 min (Table S9) face-to-face administrations were on average 1 minute shorter than remote administrations of the test (Table S10); and across diagnoses, healthy individuals completed the test 3 mins faster than cases with a diagnosis of dementia. (Table S11)

The t-test of visit 1 to visit 2 time, p=0.03. This suggests that as expected the second test takes less time to complete compared to the first test. The t-test of face to face vs remote test time taken p=0.16, suggesting there is no significant difference in time taken across the two scenarios. Whilst the mean time taken of all tests is less than 10 mins, all cases where the time taken was >15 mins occurred for dementia patients.

### Tablet use

From all the participants who completed the questionnaire (n=99), approximately 41% had no previous experience of using tablets, while ∼35% used tablets on a daily or frequent basis, the rest used tablets only a few times a week (%10) or otherwise very rarely (∼14%).

Healthy participants were more likely to be frequent users of tablets compared to MCI and Dementia patients. It is important to consider here that the sample size for healthy patients is smaller (n=15) than MCI patients (n=19), and Dementia patients (n= 54) (Figure S1).

### Smartphone use

There is a higher percentage of regular smartphone usage (∼%45) –compared to tablet–, however a similar pattern exists suggesting that those with higher levels of cognitive impairment use smartphone devices on a less regular basis. (Figure S2)

### ICA Remote operation

22 participants attempted the ICA remotely for their first visit, with only 1 participant failing to complete the ICA as they did not wish to continue. The majority were able to operate the ICA remotely without difficulty. 55% of participants found it very easy or easy to operate the device from receiving the device to starting the instructions. 19% rated the experience with medium difficulty.

Those with some previous tablet experience found it easier to operate the device compared to those who had never used tablets previously.

In the retest (7 participants conducted a remote retest), none of the participants indicated a difficult/very difficult experience in operating the ICA remotely.

### Difficulty in understanding ICA instructions

The majority of participants (∼%68) found understanding the ICA instructions either very easy or easy.

Those with cognitive impairment (particularly dementia patients) were more likely to find understanding the ICA test instructions difficult or very difficult (Figure S3).

Those with some tablet experience were more likely to find understanding the test instructions very easy or easy, and less likely to find it difficult. However a confounding factor is that healthy patients are more likely to use tablet devices.

Looking just at dementia patients, we find that within this subgroup the same pattern of easier understanding of instructions with increased tablet use exists.

On retest visits, in comparison to the first visit, a higher proportion of participants found the test instructions very easy or easy (80%), with a lower percentage indicating difficulty.

Most participants had no or little anxiety in taking the ICA. On self-reported anxiety levels, MCI and dementia participants were more likely to report more anxiety compared to other groups. 10% of MCI and dementia cases said they became very anxious, compared to none (0%) in other groups.

All participants who undertook a retest reported either the same or lower level of anxiety on the retest. There were no reports of increased anxiety on the retest.

### Difficulty in taking the ICA test itself

Overall most participants found the ICA test challenging, with less than 20% identifying the test as very easy or easy. This is to be expected as the test is designed to be challenging, there is no ceiling effect, and it is rare even for young healthy individuals to get a full score on the test.

However, after the retest the perception of participants changes, with the most common response being ‘medium’ difficulty (Figure S4).

Approximately 55% of those who did a retest indicated the second test was as difficult as their first, while 46% indicated they found the second test easier.

In the majority of cases participants were able to carry out the test without distraction, whether in face to face visits (∼85%) or remotely (∼70%). Participants were more likely to report being ‘a little distracted’ in remote tests compared to face to face tests (21% compared to 8%).

### How did the participants hold the device?

There is variation in how participants held the iPad during the test. The most common (and recommended) position was held in both hands, however ∼22% of participants placed the iPad on a stand, and ∼32% placed the iPad flat on the table.

Participants with cognitive impairment were less likely to hold the device in both hands, and more likely to place the tablet flat on the table.

However, with some tablet use, participants are more likely to hold the device in both hands. Figure S5 in the supplement shows this for dementia patients.

### Engagement and willingness to take the test again

Across all participants, a large majority (∼84%) indicated they would be willing to take the ICA again in the future. The percentage was slightly lower for those with cognitive impairment (∼78%).

The percentage of those willing to take the ICA again increased to over 90% for those who also completed the retest.

## Discussion

This study, conducted as part of the ADePT project, has provided quantitative and qualitative evidence on the effectiveness and usability of the ICA in real world clinical environments. In other studies, such as Kalafatis et al. 2021, the sensitivity of the ICA in detecting cognitive impairment in MCI and mild AD patients was investigated, as well as the convergent validity of the ICA with reference cognitive tests such as MoCA and ACE-III. This study focused on comparison of the ICA classification with GP referral, and ultimately the specialist diagnosis from the memory clinics. The ICA was tested in a wider range of patients who had been referred to memory clinics, including those with more progressed dementia, those with non-dementia conditions, and a number of inconclusive cases.

The primary outcome of the study compared GP referral with specialist diagnosis of MCI/dementia. Of those the GP referred to memory clinics, 78% were necessary referrals, with ∼22% unnecessary referrals, or patients who should have been referred to other services as they had disorders other than MCI/dementia. In the same population the ICA was able to correctly identify cognitive impairment in ∼90% of patients, with approximately 9% of patients being false negatives. From the subset of unnecessary GP referrals, the ICA classified ∼72% of those as not having cognitive impairment, suggesting that these unnecessary referrals may not have been made if the ICA was in use.

The proportion of necessary GP referral in our work is higher than what has been found in other studies like Creavin, et al. (2021), where 56% was reported. It is likely that the GPs referrals assessed for the purpose of this study had a special interest in Dementia diagnosis, thus the higher rate of accurate referrals. The sample size here is less than 100 participants from four sites in the UK, therefore GP referral accuracy from larger studies should also be considered. However the GP referral precision obtained from this study aligns with the values obtained from Sussex audit report for 2020-2021, which indicated there was no diagnosis of cognitive impairment in only 8% of cases from those who presented at the memory clinic.

Due to the challenges in recruiting patients from various GP sites, a main limitation of this study is that only those who were referred by GPs have been investigated. Therefore the results from this study cannot be used to determine the false negative rate, i.e. those who did have cognitive impairment but were not referred by their GP. We seek to address this in future studies where the ICA will also be deployed in primary care.

In response to the COVID-19 pandemic, and the move towards remote assessments, the study protocol was amended to allow remote administration of the ICA. Twenty-two participants attempted the ICA remotely, with only one participant not wishing to continue. Most patients found the remote operation of cognICA very easy or easy, however those with no prior tablet experience were more likely to have difficulty. In the majority of cases participants were able to carry out remote tests without distraction, however participants were more likely to report slight distraction in remote tests compared to face to face tests.

Overall the usability results show that most participants were able to understand the ICA instructions and complete the test, even though they found the rapid visual categorisation task itself challenging. Patient’s usability experience was found to be related to previous familiarity with phones and tablets; those with some prior experience found it easier to understand the instructions and to operate the device. They were also more likely to hold the device in both hands rather than placing it flat on the table. There were also differences in perception amongst patients based on their clinical diagnosis. As expected those with MCI, dementia or a non-dementia condition were more likely to have difficulties, and to feel slight or mild anxiety in taking the ICA.

Those who took a retest reported an improved overall usability experience. They found the instructions easier to understand, experienced less or the same level of anxiety, found the test easier or the same as before and found operating the device in remote assessments easier than the first visit. Both after the first, and second visit, a significant majority of patients indicated they would be willing to take the ICA again in the future.

Some patients did not manage to complete the test, in particular those with dementia or more severe cognitive impairment. An update to the ICA software which allowed participants to seamlessly progress from the pre-test demo and trial images to the main test, as well as the introduction of a new instruction video helped to increase the completion rate. For dementia patients the increase was from 78% in cognICA 1.2 to 89% in cognICA 1.3. It is important to note that ICA is particularly designed to detect and monitor subtle cognitive impairments or changes to cognitive performance in earlier stages of the disease, where there is an unmet need in clinical practice.

The ICA correlation with cognitive tests, taken in memory clinics and GP settings was also calculated, where there was at most 12 months between the assessments. The ICA index had a Pearson correlation of 0.57 (p<0.000) with ACE-III, 0.77 (p<0.002) with MoCA which were taken as part of the memory clinic assessment. The ICA had a lower correlation with the more rudimentary assessments taken in the GP setting, e.g. a correlation of 0.37 (p<0.18) with GPCOG.

## Conclusions

The results from this study demonstrate the potential of the ICA as a screening tool, which can be used to support accurate referrals from primary care settings, along with the work conducted in memory clinics and in secondary care. The study investigated a cohort of patients aged 55-90, who were recruited at NHS memory services. The aim of the study was to gather real-world evidence of the use of the ICA in memory clinics and compare the performance of the ICA versus GP referrals.

The ICA test has proven to be a consistent method of detection of cognitive impairment, with an initial test and a retest being strongly, positively correlated (r=0.88, p<0.000) and a test retest prediction agreement of 87.5%. Furthermore, the ICA and more traditional tests, like ACE-III or MoCA, share a percent agreement in the range of 70-90%. Analysis demonstrated that, if adopted, the ICA would have reduced unnecessary referrals from GPs. Paired with a faster delivery time (approximately 5-10 minutes, compared to about 30 minutes for a traditional test like ACE-III), the ICA can prove an effective tool in the assessment of cognitive impairment (Shore et al. 2022).

Most participants across clinical diagnoses could complete the test and understand the ICA instructions, even though about half of the cohort would very rarely or never use a tablet. A third of the cohort found the test challenging, but patients’ usability experience improved during the retest. Furthermore, the introduction of an updated version of the cognICA software (with pre-test demo and trial images available to users) has proved to increment user engagement and completion rate, demonstrating the potential of this tool.

## Data Availability

All data produced in the present work are contained in the manuscript.

