## Supplementary materials for "The Use of the Integrated Cognitive Assessment (ICA) to Improve the Efficiency of Primary Care Referrals to Memory Services in the Accelerating Dementia Pathway Technologies (ADePT) Study"

### Appendix

#### A1 ICA correlation with cognitive tests

In the ADePT study recent and historical cognitive and functional test scores, taken in memory clinics and GP settings are captured. A summary of all of the cognitive test results is shown in Table S1.

***Table S1.*** *Cognitive test scores taken by participants in memory clinics and GP settings.*

| | | Score | | | | | Days difference with ICA test | | | | |
|---|---|---|---|---|---|---|---|---|---|---|---|
| | | Number of data points | mean | std | min | max | Number of data points | mean | std | min | max |
| category | name | | | | | | | | | | |
| ACE_historical | ACE-III | 10 | 87.7 | 7.1 | 71 | 97 | 10 | 1036 | 680 | 103 | 2355 |
| ACE_memory_clinic | ACE-III | 68 | 76.1 | 16.8 | 13 | 96 | 68 | 40 | 54 | 8 | 310 |
| AMTS_GP | AMTS | 1 | 4.0 | Na | 4 | 4 | 1 | 111 | Na | 111 | 111 |
| BADLS | BADLS | 8 | 7.0 | 6.8 | 0 | 20 | 6 | 244 | 370 | 22 | 989 |
| GPCOG_GP | GPCOG | 20 | 6.9 | 3.7 | 0 | 12 | 19 | 231 | 493 | 36 | 2157 |
| MCOG_GP | MCOG | 1 | 3.0 | Na | 3 | 3 | 1 | 163 | Na | 163 | 163 |
| MMSE_GP | MMSE | 6 | 23.8 | 8.6 | 8 | 30 | 5 | 290 | 293 | 68 | 776 |
| MOCA_BLIND_memory_clinic | MoCA-BLIND | 14 | 11.4 | 3.7 | 3 | 18 | 14 | 28 | 7 | 16 | 41 |
| MOCA_historical | MoCA | 9 | 24.2 | 4.3 | 16 | 28 | 9 | 744 | 618 | 69 | 1977 |
| MOCA_memory_clinic | MoCA | 12 | 24.4 | 6.3 | 14 | 30 | 12 | 67 | 59 | 0 | 182 |
| OTHER_FUN_TEST | CBI-R | 1 | 13.0 | Na | 13 | 13 | 1 | 27 | Na | 27 | 27 |
| | CBI-R (for the carer) | 1 | 24.0 | Na | 24 | 24 | 1 | 118 | Na | 118 | 118 |
| | Cambridge Behavioural Inventory Revised | 1 | 17.0 | Na | 17 | 17 | 1 | 260 | Na | 260 | 260 |
| OTHER_TEST_GP | MoCA | 1 | 16.0 | Na | 16 | 16 | 1 | 69 | Na | 69 | 69 |
| OTHER_memory_clinic | Other | 1 | 96.0 | Na | 96 | 96 | 1 | 66 | Na | 66 | 66 |
| Other_test_historical | 6CIT | 2 | 4.5 | 3.5 | 2 | 7 | 2 | 982 | 427 | 680 | 1284 |
| | ACE-R | 1 | 85.0 | Na | 85 | 85 | 1 | 1725 | Na | 1725 | 1725 |
| | Abbreviated Mental Test Score | 1 | 4.0 | Na | 4 | 4 | 1 | 111 | Na | 111 | 111 |
| | CAMCOG | 1 | 93.0 | Na | 93 | 93 | 1 | 1629 | Na | 1629 | 1629 |
| | GPCOG | 1 | 5.0 | Na | 5 | 5 | 1 | 40 | Na | 40 | 40 |
| | Test Your Memory | 1 | 48.0 | Na | 48 | 48 | 1 | 123 | Na | 123 | 123 |
| SCIT_GP | SCIT | 21 | 7.8 | 6.5 | 0 | 22 | 21 | 264 | 372 | 52 | 1336 |
| TYM_GP | TYM | 1 | 48.0 | Na | 48 | 48 | 1 | 123 | Na | 123 | 123 |

We calculated the correlation between ICA Index with cognitive tests taken in memory clinics and GP settings where there was at most 12 months difference in date between the tests. Results are included where there were at least 10 data points for the correlation calculation. The ICA Pearson correlation of 0.57 with ACE-III, 0.77 with MoCA, and 0.37 with GPCOG establishes convergent validity with these tests. The ICA has a higher correlation with the more comprehensive cognitive assessments such as MoCA and ACE than with shorter assessments such as GPCOG.

***Table S2.*** *The correlation between cognitive tests scores and ICA Index where there was at most 12 months difference in date between the two tests, and at least 10 data points*

| **metric1** | **metric2** | **number_points** | **pearson correlation** | **pearson p-value** | **spearman correlation** | **Spearman p-value** | **number_points** | **mean_days_between_the_tests** | **std_days_between_the_tests** | **max_days_between_the_tests** | **min_days_between_the_tests** | **median_num_days** |
|---|---|---|---|---|---|---|---|---|---|---|---|---|
| ICA_test_1 | ACE-III | 64 | 0.57 | 0.0000 | 0.58 | 0.0000 | 64.0 | 44.3 | 62 | 310 | 8 | 26.5 |
| ICA_test_1 | ACE-III_attention | 63 | 0.48 | 0.0001 | 0.47 | 0.0001 | 63.0 | 40.2 | 53 | 310 | 8 | 26 |
| ICA_test_1 | ACE-III_fluency | 63 | 0.50 | 0.0000 | 0.50 | 0.0000 | 63.0 | 40.2 | 53 | 310 | 8 | 26 |
| ICA_test_1 | ACE-III_language | 63 | 0.49 | 0.0001 | 0.50 | 0.0000 | 63.0 | 40.2 | 53 | 310 | 8 | 26 |
| ICA_test_1 | ACE-III_memory | 63 | 0.47 | 0.0001 | 0.43 | 0.0004 | 63.0 | 40.2 | 53 | 310 | 8 | 26 |
| ICA_test_1 | ACE-III_visuospatial | 63 | 0.49 | 0.0000 | 0.47 | 0.0001 | 63.0 | 40.2 | 53 | 310 | 8 | 26 |
| ICA_test_1 | GPCOG | 14 | 0.37 | 0.1879 | 0.32 | 0.2669 | 14.0 | 82.1 | 46 | 210 | 36 | 67 |
| ICA_test_1 | MoCA | 13 | 0.77 | 0.0021 | 0.83 | 0.0004 | 13.0 | 96.7 | 78 | 298 | 21 | 101 |
| ICA_test_1 | MoCA-BLIND | 12 | 0.09 | 0.7865 | 0.16 | 0.6184 | 12.0 | 28.4 | 7 | 41 | 17 | 28.5 |
| ICA_test_1 | SCIT | 15 | -0.07 | 0.7978 | 0.00 | 0.9949 | 15.0 | 117.1 | 50 | 246 | 52 | 110 |

ICA and ACE-III predictions

***Table S3.*** *Details on the patients who had different predictions from ACE-III and ICA*

| Clinical diagnosis | Non-dementia condition | Age | Education years | ICA Index | ICA score | ACE score | ACE Attention | ACE Memory | ACE Fluency | ACE Language | ACE Visuospatial |
|---|---|---|---|---|---|---|---|---|---|---|---|
| Dementia | | 76 | 10 | 69 | 74 | 83 | 17 | 16 | 11 | 24 | 15 |
| Dementia | | 73 | 11 | 53 | 1 | 90 | 16 | 24 | 12 | 26 | 12 |
| Dementia | | 76 | 18 | 68 | 92 | 77 | 13 | 14 | 10 | 25 | 15 |
| Dementia | | 73 | 20 | 66 | 81 | 84 | 16 | 17 | 9 | 26 | 16 |
| Healthy | | 62 | 24 | 59 | 47 | 93 | 18 | 23 | 10 | 26 | 16 |
| Healthy | | 71 | 14 | 54 | 1 | 94 | 16 | 25 | 12 | 25 | 16 |
| Inconclusive | | 73 | 17 | 31 | | 79 | 17 | 14 | 8 | 24 | 16 |
| MCI | | 74 | 11 | 48 | 3 | 88 | 16 | 21 | 9 | 26 | 16 |
| MCI | | 73 | 20 | 52 | 16 | 96 | 16 | 25 | 13 | 26 | 16 |
| MCI | | 68 | 15 | 51 | 4 | 88 | 18 | 22 | 10 | 24 | 14 |
| MCI | | 82 | 11 | 77 | 93 | 84 | 18 | 16 | 9 | 25 | 16 |
| MCI | | 71 | 11 | 21 | 1 | 88 | 17 | 22 | 9 | 24 | 16 |
| Non-dementia conditio | Z71.7 Age associated | 82 | 10 | 47.1808 | 1.047262301 | 96 | 18 | 23 | 13 | 26 | 16 |

| | | | | | | | | | | | |
|---|---|---|---|---|---|---|---|---|---|---|---|
| n | memory decline | | | | | | | | | | |
| Non-dementia condition | Cerebral amyloid angiopathy (CAA) | 81 | 15 | 50.4202 | 1.219038622 | 89 | 15 | 25 | 9 | 26 | 14 |

ICA - MoCA Predictions

***Table S4***. *Instances of different prediction between MoCA and ICA*

| **Clinical diagnosis** | **Non-dementia condition** | **Age** | **Education years** | **ICA Index** | **ICA score** | **AI prediction** | **MoCA score** | **MOCA prediction** |
|---|---|---|---|---|---|---|---|---|
| Healthy | | 81 | 11 | 47.0 | 2.0 | Impaired | 26 | Healthy |
| Inconclusive | | 77 | 16 | 52.0 | 6.6 | Impaired | 28 | Healthy |
| MCI | | 80 | 18 | 56.2 | 4.4 | Impaired | 28 | Healthy |

ICA - MoCA Blind

***Table S5.*** *Instances of different prediction between MoCA Blind and ICA*

| Clinical diagnosis | Non-dementia condition | Age | Education years | ICA Index | ICA score | AI prediction | MoCA Blind score | MOCA_BLIND_memory_clinic_prediction |
|---|---|---|---|---|---|---|---|---|
| MCI | | 73 | 10 | 43.3 | 1.0 | Impaired | 18 | Healthy |

ICA - GPCOG

***Table S6.*** *Instances of different prediction between GPCOG and ICA*

| Clinical diagnosis | Non-dementia condition | Age | Education years | ICA Index | ICA score | AI prediction | GPCOG score | GPCOG_GP_prediction |
|---|---|---|---|---|---|---|---|---|
| Dementia | | 73 | 11 | 52.9 | 1.1 | Impaired | 9 | Healthy |
| Dementia | | 76 | 18 | 68.0 | 92.3 | Healthy | 5 | Further Investigate |
| Dementia | | 63 | 13 | 29.8 | 1.0 | Impaired | 6 | Further Investigate |
| Dementia | | 84 | 10 | 37.6 | 1.0 | Impaired | 9 | Healthy |
| Dementia | | 77 | 10 | 40.5 | 1.0 | Impaired | 9 | Healthy |
| Dementia | | 73 | 20 | 65.8 | 80.9 | Healthy | 7 | Further Investigate |
| Healthy | | 81 | 11 | 47.0 | 2.0 | Impaired | 11 | Healthy |
| MCI | | 79 | 10 | 37.3 | 1.0 | Impaired | 10 | Healthy |

ICA - 6CIT

***Table S7.*** *Instances of different prediction between 6CIT and ICA*

| Clinical diagnosis | Non-dementia condition | Age | Education years | ICA Index | ICA score | AI prediction | SCIT score | SCIT_cut_off_prediction |
|---|---|---|---|---|---|---|---|---|
| Dementia | | 78 | 10 | 45.0 | 1.1 | Impaired | 3 | Healthy |
| Dementia | | 88 | 11 | 9.2 | 1.0 | Impaired | 2 | Healthy |

| | | | | | | | | |
|---|---|---|---|---|---|---|---|---|
| Dementia | | 76 | 10 | 69.2 | 74.0 | Healthy | 8 | Impaired |
| Dementia | | 84 | 15 | 58.0 | 1.2 | Impaired | 4 | Healthy |
| Inconclusive | | 85 | 11 | 7.0 | 1.0 | Impaired | 4 | Healthy |
| Inconclusive | | 65 | 11 | 68.9 | 96.1 | Healthy | 11 | Impaired |

#### A2 ICA Usability

ICA Completion Time

***Table S8.*** *Overall completion time of the ICA test*

| Participants | Number of participants | Mean time (mins) | std | min time (mins) | 25th percentile time (mins) | Median | 75th percentile time (mins) | max time (mins) |
|---|---|---|---|---|---|---|---|---|
| All completed ICA tests (visit 1 and 2) | 106 | 8.5 | 3.7 | 2.0 | 6.0 | 8.0 | 10.0 | 23.0 |

***Table S9.*** *Time taken in visit 1 compared to visit 2*

| Participants | Number of participants | Mean time (mins) | std | min time (mins) | 25th percentile time (mins) | Median | 75th percentile time (mins) | max time (mins) |
|---|---|---|---|---|---|---|---|---|
| Visit 1 ICA tests | 80 | 9.0 | 4.0 | 2.0 | 6.0 | 8.0 | 11.0 | 23.0 |
| Visit 2 ICA tests | 24 | 7.2 | 2.4 | 4.0 | 5.0 | 7.0 | 8.8 | 15.0 |

***Table S10.*** *Time taken in face to face visits compared to remote visits*

| Participants | Number of participants | Mean time (mins) | std | min time (mins) | 25th percentile time (mins) | Median | 75th percentile time (mins) | max time (mins) |
|---|---|---|---|---|---|---|---|---|
| **Face to Face** | 79 | 8.3 | 3.9 | 2.0 | 5.0 | 8.0 | 10.0 | 23.0 |
| **Remote** | 27 | 9.4 | 3.1 | 5.0 | 7.0 | 9.0 | 10.5 | 17.0 |

***Table S11.*** *Time taken for different diagnosis groups*

| Participants | Number of participants | Mean time (mins) | std | min time (mins) | 25th percentile time (mins) | Median | 75th percentile time (mins) | max time (mins) |
|---|---|---|---|---|---|---|---|---|
| **Dementia** | 58 | 9.5 | 4.2 | 2.0 | 6.3 | 9.0 | 11.8 | 23.0 |
| **MCI** | 22 | 8.0 | 2.3 | 4.0 | 6.3 | 8.0 | 9.8 | 12.0 |
| **Healthy** | 17 | 6.6 | 1.8 | 4.0 | 5.0 | 6.0 | 8.0 | 10.0 |
| **Non-dementia condition** | 5 | 6.2 | 2.2 | 5.0 | 5.0 | 5.0 | 6.0 | 10.0 |
| **Inconclusive** | 4 | 9.3 | 5.3 | 5.0 | 6.5 | 7.5 | 10.3 | 17.0 |

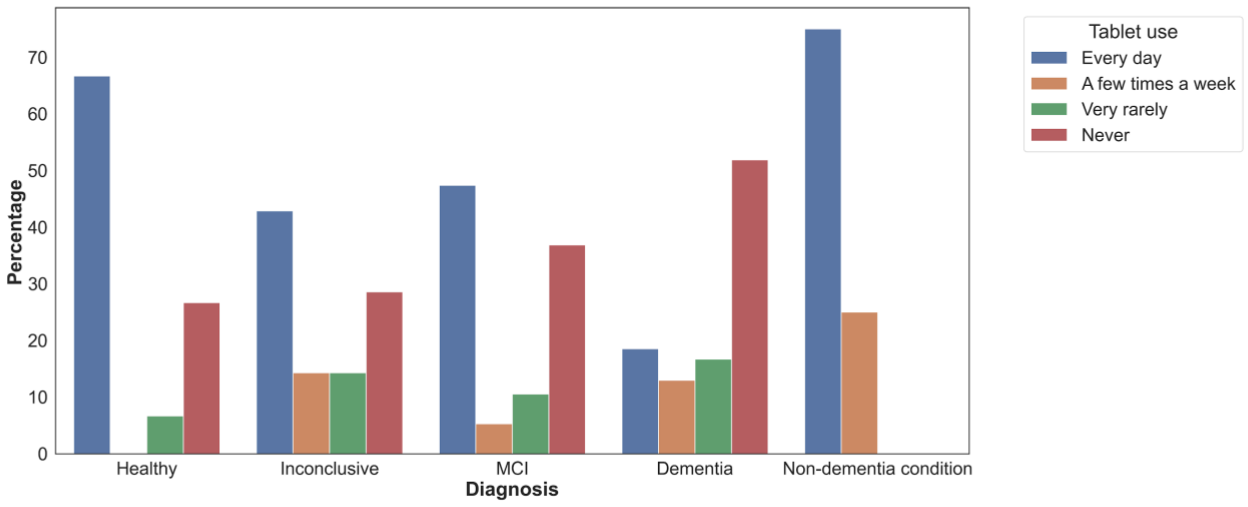


***Figure S1.*** *Breakdown of healthy, MCI and Dementia patients relative to their tablet use*

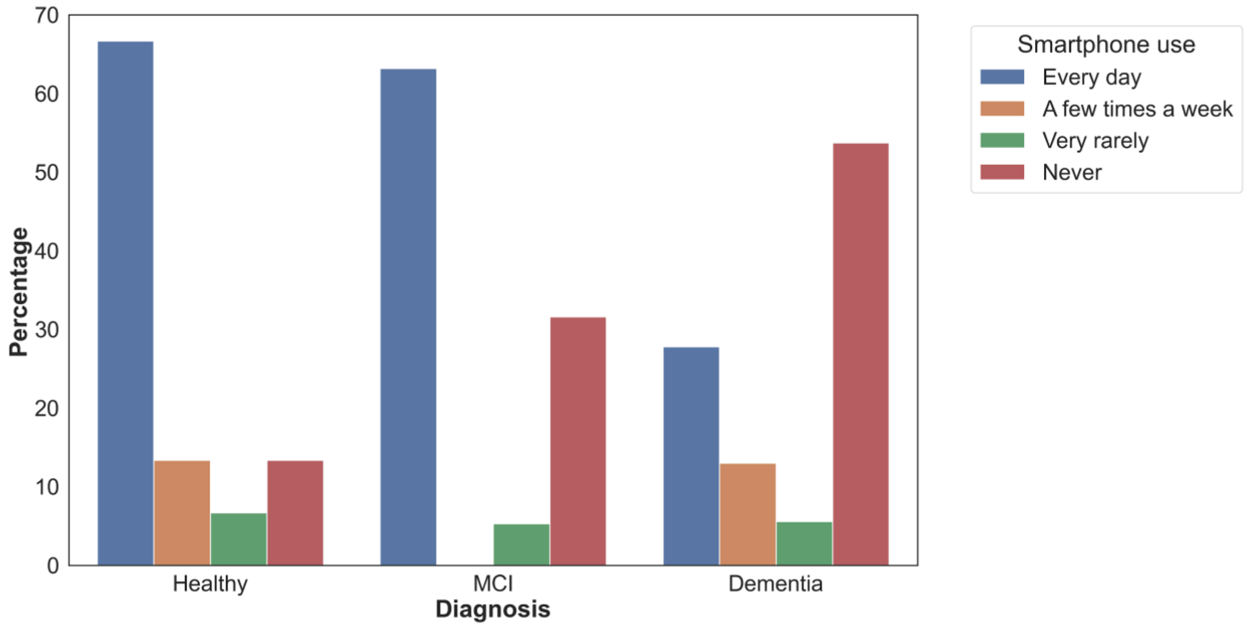


***Figure S2***. *Breakdown of healthy, MCI and Dementia patients relative to their smartphone use*

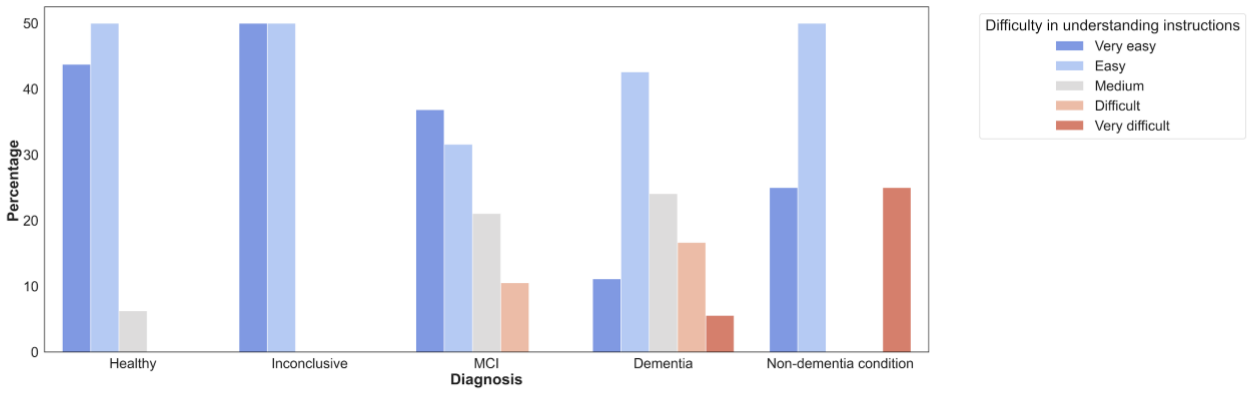


***Figure S3.*** *Difficulty in understanding the ICA test instructions, broken down by participants*

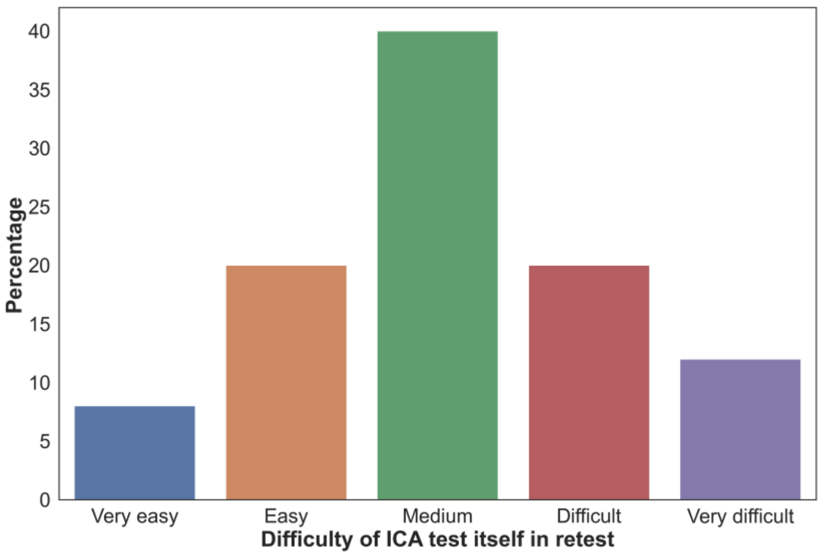


**Figure S4.** How difficult participants found the ICA test itself on their retest

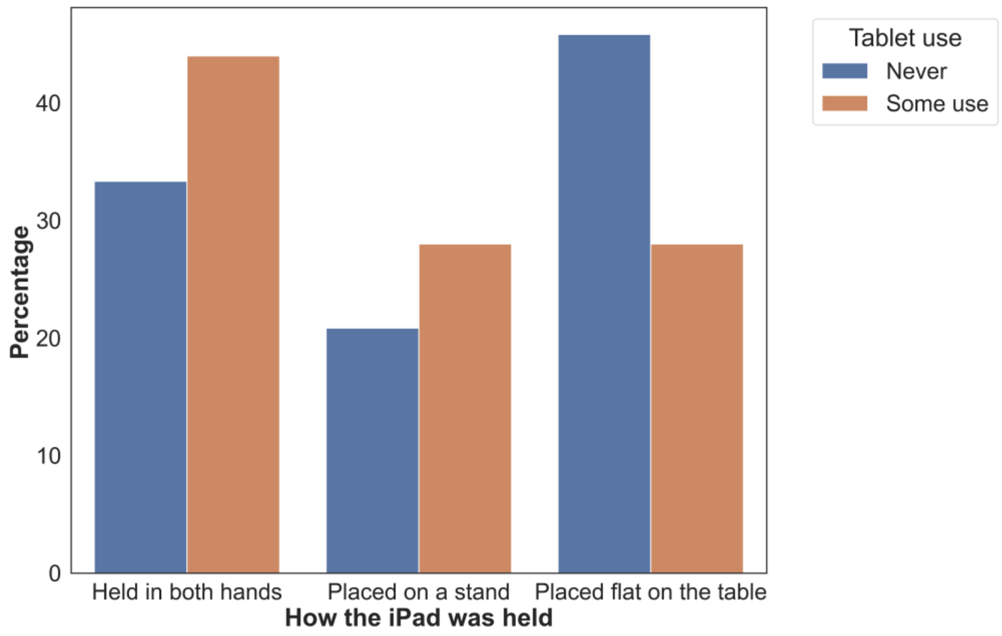


***Figure S5.*** *How dementia participants positioned the iPad during the test broken down by their previous experience with tablets*

##### A3 Cognitive health questionnaire

The responses to each of the questions on the cognitive health questionnaire, broken down by clinical diagnosis is shown in Figures below.

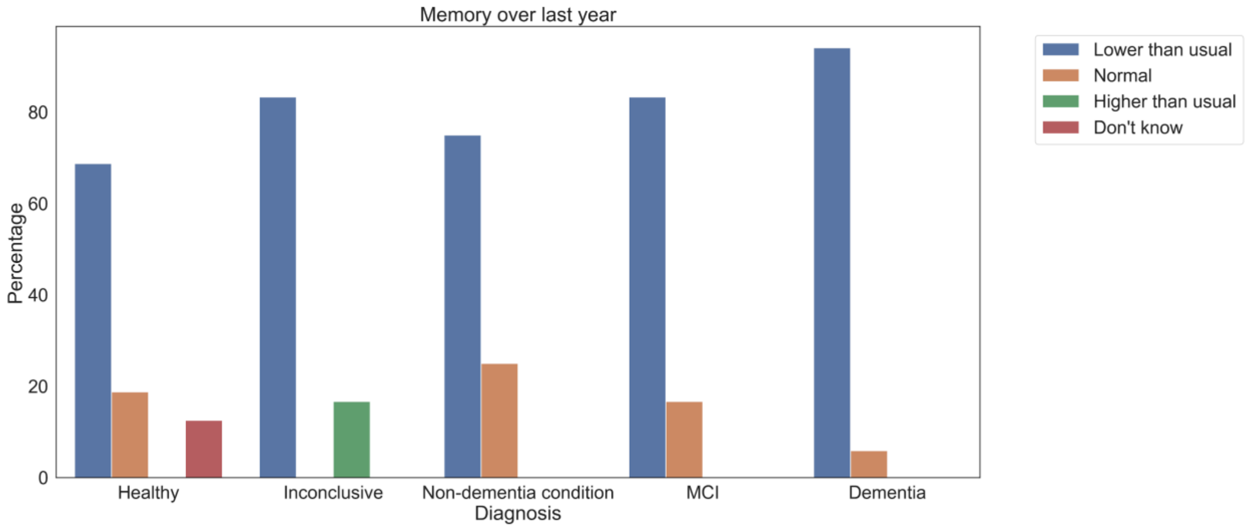


***Figure S6.*** *Change in memory over the last year*

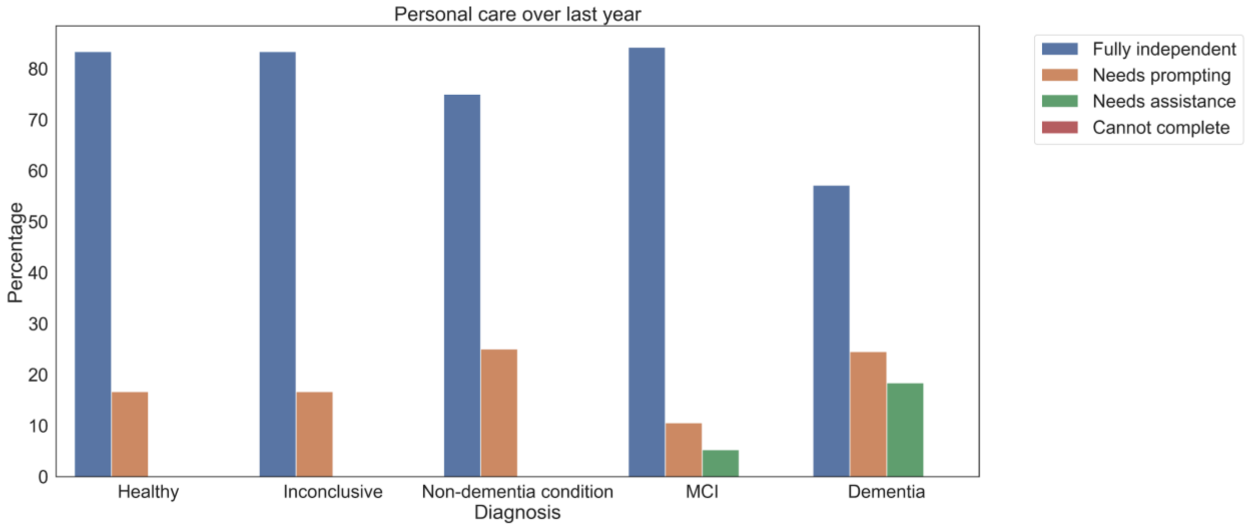


***Figure S7.*** *Personal care over the last year*

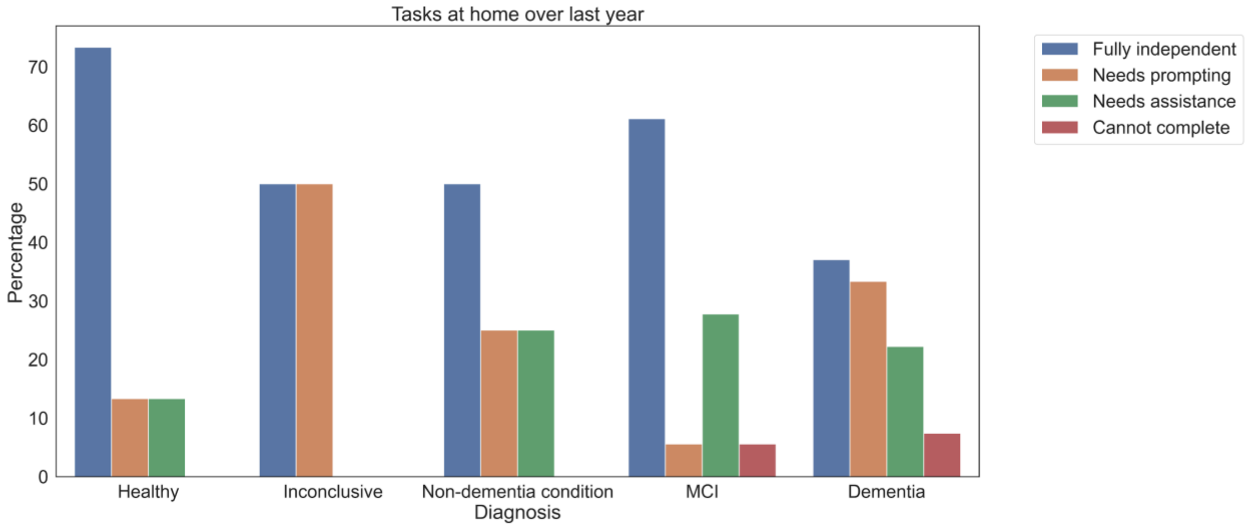


***Figure S8.*** *Tasks at home over the last year*

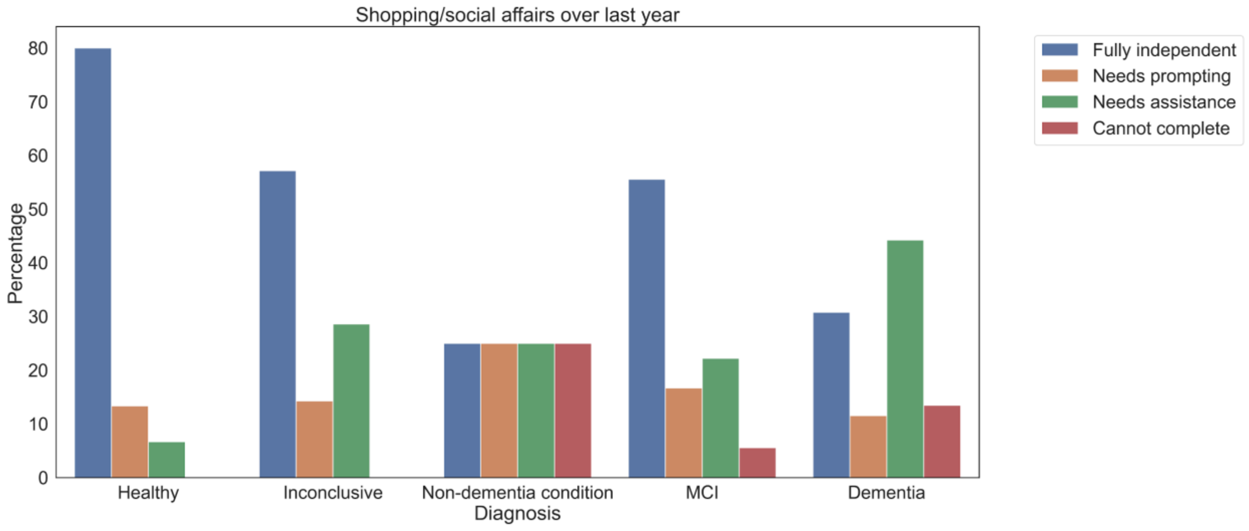


***Figure S9.*** *Shopping/social affairs over the last year*

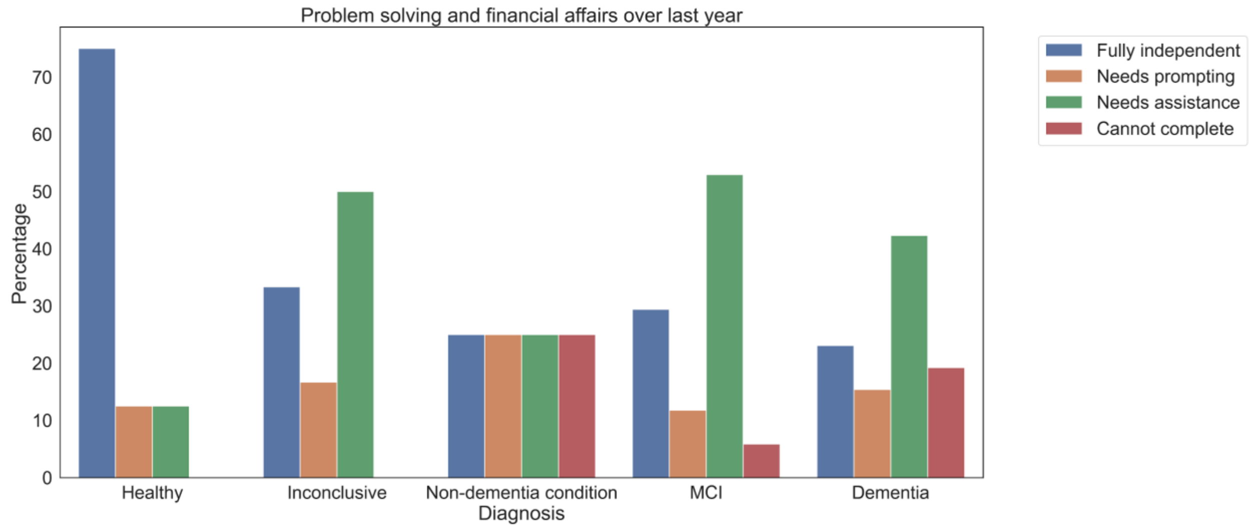


***Figure S10**. Problem solving and financial affairs over the last year*

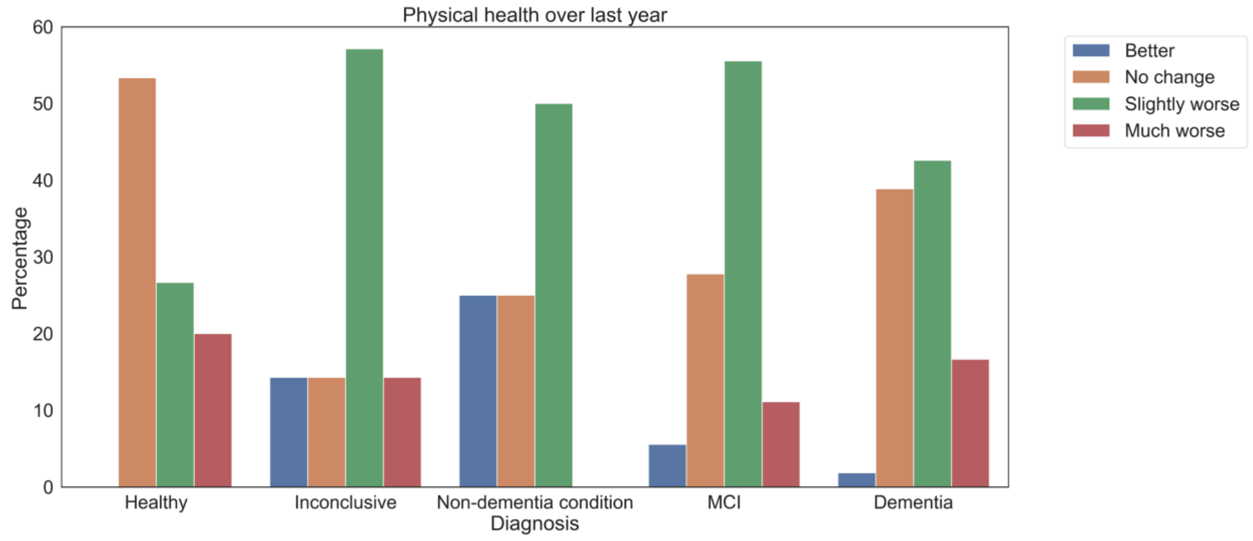


***Figure S11.** Physical health over the last year*

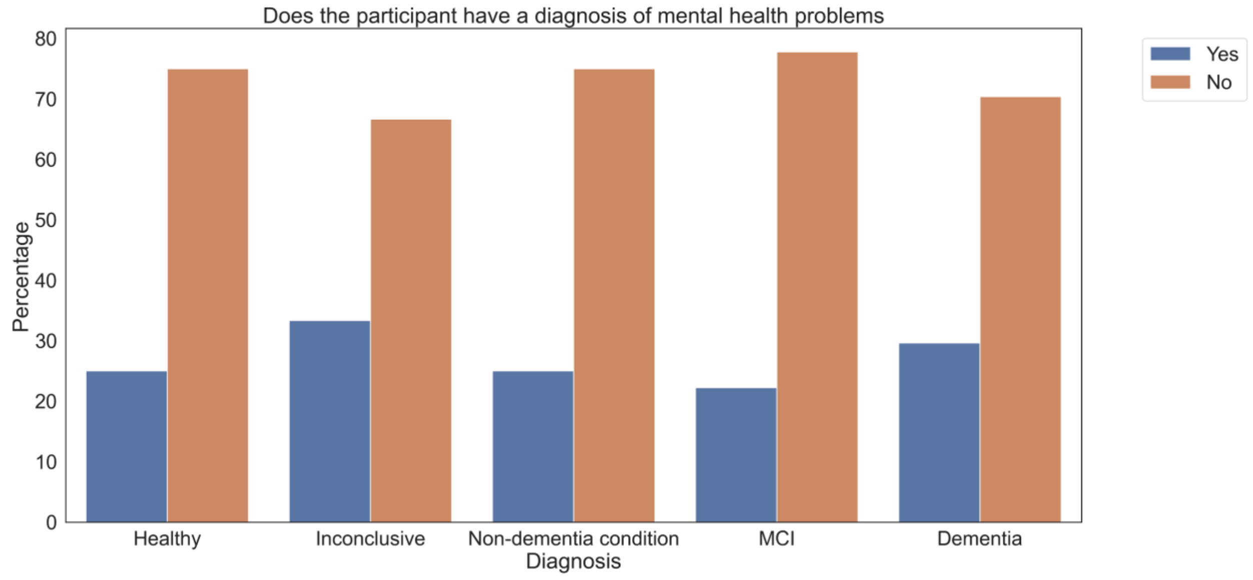


***Figure S12**. History of mental health problems*

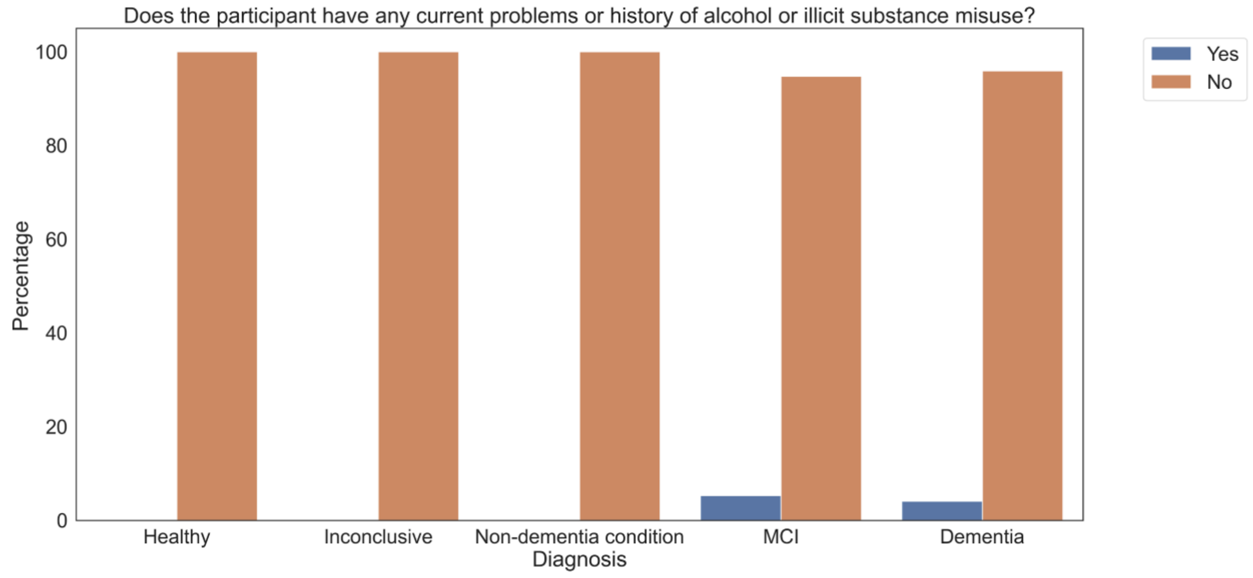


***Figure S13.** History of alcohol or illicit substance misuse*

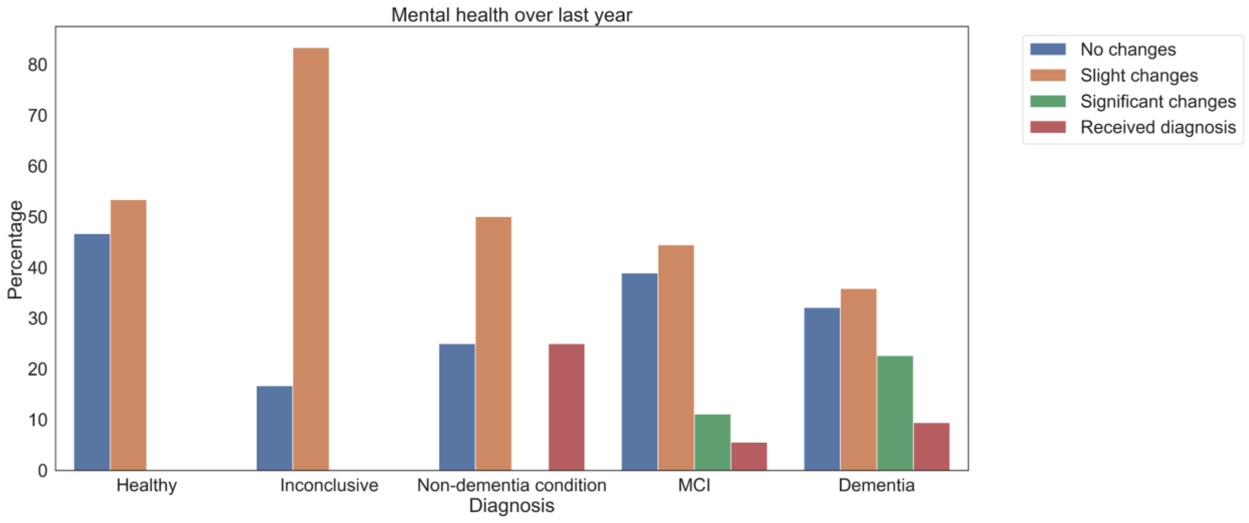


***Figure S14.*** *Mental health over last year*

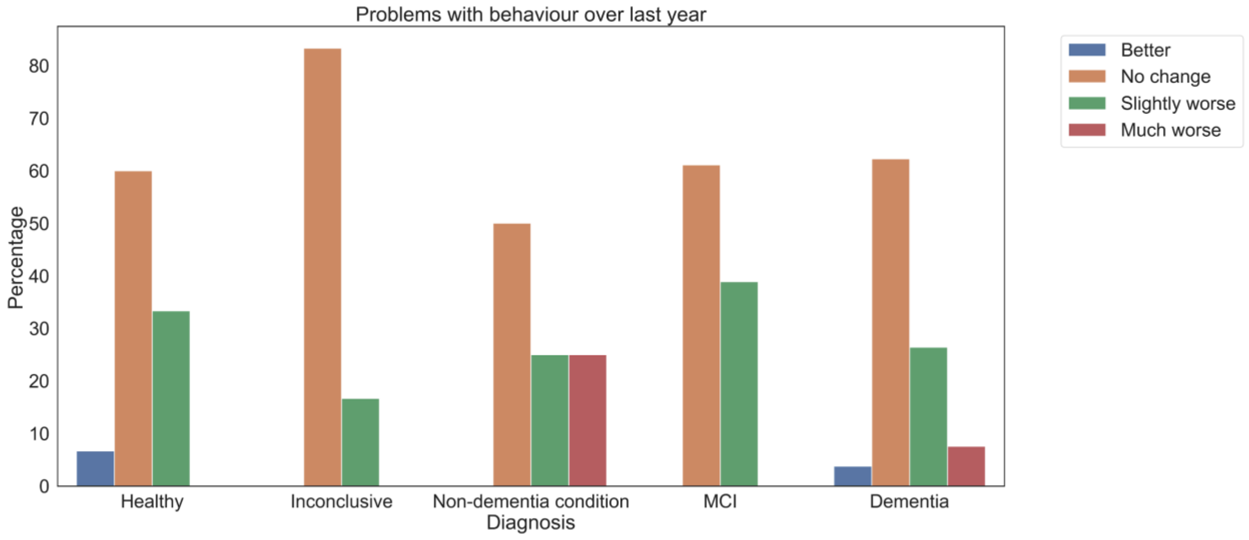


***Figure S15.*** *Problems with behaviour over last year*
